# Extensive cryptic circulation sustains mpox among men who have sex with men

**DOI:** 10.1101/2025.08.09.25333368

**Authors:** Joseph A. Lewnard, Miguel I. Paredes, Matan Yechezkel, Gregg S. Davis, Vennis Hong, Jessica Skela, Utsav Pandey, Noah T. Parker, Lauren C. Granskog, Magdalena E. Pomichowski, Iris Anne C. Reyes, Isabel Rodriguez-Barraquer, Nicola F. Müller, Sara Y. Tartof

## Abstract

Sporadic cases of mpox continue to be notified among men who have sex with men (MSM), with most lacking identifiable transmission links. To resolve underlying dynamics, we tested prospectively for monkeypox virus (MPXV) in rectal swabs from a cohort of MSM in Los Angeles, whom we monitored concurrently for clinical mpox diagnoses during summer, 2024. Here we show that MPXV infections exceeded reported mpox cases by a 33-fold margin (95% confidence interval: 16-68), revealing incidence rates of MPXV infection comparable to *Neisseria gonorrhoeae* and *Chlamydia trachomatis* among MSM. Independent estimates derived from MPXV phylogenetic reconstruction and a meta-analysis of surveillance studies corroborated this extensive under-reporting. We estimate that undiagnosed infections must cause at least 31-44% of all transmission to explain observed MPXV phylogenies; under realistic modeling assumptions, this proportion rises to 61-94%. Contrary to current guidance, our findings suggest MPXV is prevalent among MSM, predominantly causes subclinical infection, and is sustained in circulation by cryptic transmission. The feasibility of current mpox elimination targets merits assessment in light of prevalent and extensively under-reported infections.

---

Since 2022, clade IIb monkeypox virus (MPXV) has continued to spread sexually among men who have sex with men (MSM) and other sexual- and gender-minority populations in countries where it was previously non-endemic.^1^ Clinically, mpox is associated with a prodrome characterized by fever, chills, myalgia, and lymphadenopathy, followed by painful cutaneous or mucosal lesions, often localized to sites of sexual exposure.^2^ Current guidance from the US Centers for Disease Control & Prevention (CDC) and European Centre for Disease Prevention & Control (ECDC) stipulates that subclinical or asymptomatic infections are uncommon.^3,4^ Accordingly, declines in reported case counts to sporadic detections in most countries have been interpreted as evidence of successful containment,^5^ with ECDC and the World Health Organization (WHO) targeting elimination of human-to-human transmission by 2027 in countries with outbreaks driven by sexual transmission.^6,7^

However, several observations have suggested cryptic transmission that may undermine this objective. Although numerous health agencies recommend contact tracing as a strategy to interrupt transmission,^8–11^ >90% of mpox cases have lacked epidemiological links to prior cases, and 67-85% have reported no known exposure to individuals with mpox or mpox-like symptoms.^2,12–15^ Individuals shed replication-competent MPXV before symptoms onset and during subclinical infection,^16–18^ and pre-symptomatic transmission is widely documented.^2,19–21^ In contrast to current guidance,^3^ these findings imply that contact with lesions is not required for MPXV spread. Quantifying the proportion of individuals that become symptomatic and receive mpox diagnoses could clarify the epidemiological importance of subclinical MPXV infections. However, ascertainment ratios for clade IIb MPXV remain unknown.

To resolve this uncertainty, we launched a prospective surveillance study among Los Angeles MSM receiving healthcare from Kaiser Permanente Southern California (KPSC). The KPSC healthcare system provides comprehensive, integrated care to members; electronic health records (EHRs) record all diagnoses, clinical notes, laboratory tests, procedures, immunizations, and pharmacy prescription fills. Insurance reimbursement data capture out-of-network care, and history of vaccination with JYNNEOS (modified *Vaccinia* Ankara) is automatically reconciled with the California Immunization Registry, to which providers statewide are required to report all vaccine administrations. Our study cohort comprised 7,930 males aged 16-52 years on May 1, 2024, with ≥1 year of prior enrollment and ≥1 anorectal test for *Chlamydia trachomatis* or *Neisseria gonorrhoeae* infection (an indication reserved for MSM). From May 29 to November 13, 2024, we tested eligible cohort members’ remnant anorectal specimens collected for *C. trachomatis*/*N. gonorrhoeae* screening—which sexually-active MSM are recommended to receive as often as every 3-6 months—for MPXV DNA via real-time polymerase chain reaction (PCR), while concurrently monitoring for mpox diagnoses in any clinical care setting.

## Subclinical MPXV infections vastly outnumber diagnosed cases

The incidence rate of laboratory-confirmed mpox diagnoses identified through standard-of-care testing in clinical settings was 0.61 (95% confidence interval: 0.27-0.77) cases per 100 person-years at risk among cohort members (*n*=15 cases) in analyses that balanced demographic and clinical characteristics of individuals who received or did not receive screening tests through inverse propensity weighting (**Figure S1**; **Table S1**). Accounting solely for virus shedding among diagnosed cases, we expected a weighted MPXV prevalence of 0.035% in anorectal swabs (**Figure 1A**; **Table S2**; **Figure S2**). In contrast, testing 1,190 remnant anorectal specimens from 1,054 individuals, we identified 7 positive results among 6 individuals, yielding markedly higher estimates of weighted prevalence (0.91% [0.40-1.6%] and incidence (20 [8.5-48] infections per 100 person-years; see **Methods** for description of quantitative polymerase chain reaction [qPCR] testing procedures). All positive specimens had cycle threshold (c_*T*_) values below the validated detection limit for both an MPXV-specific probe and a non-Variola *Orthopoxvirus* (NVAR) probe. Within each specimen, c_*T*_ values were similar for the MPXV-specific and NVAR probes (range in absolute differences: 0.5-0.9; **Table S3**), indicating closer alignment in quantitative results than expected by chance (two-sided bootstrap *p*<0.0001).

**Figure 1:**
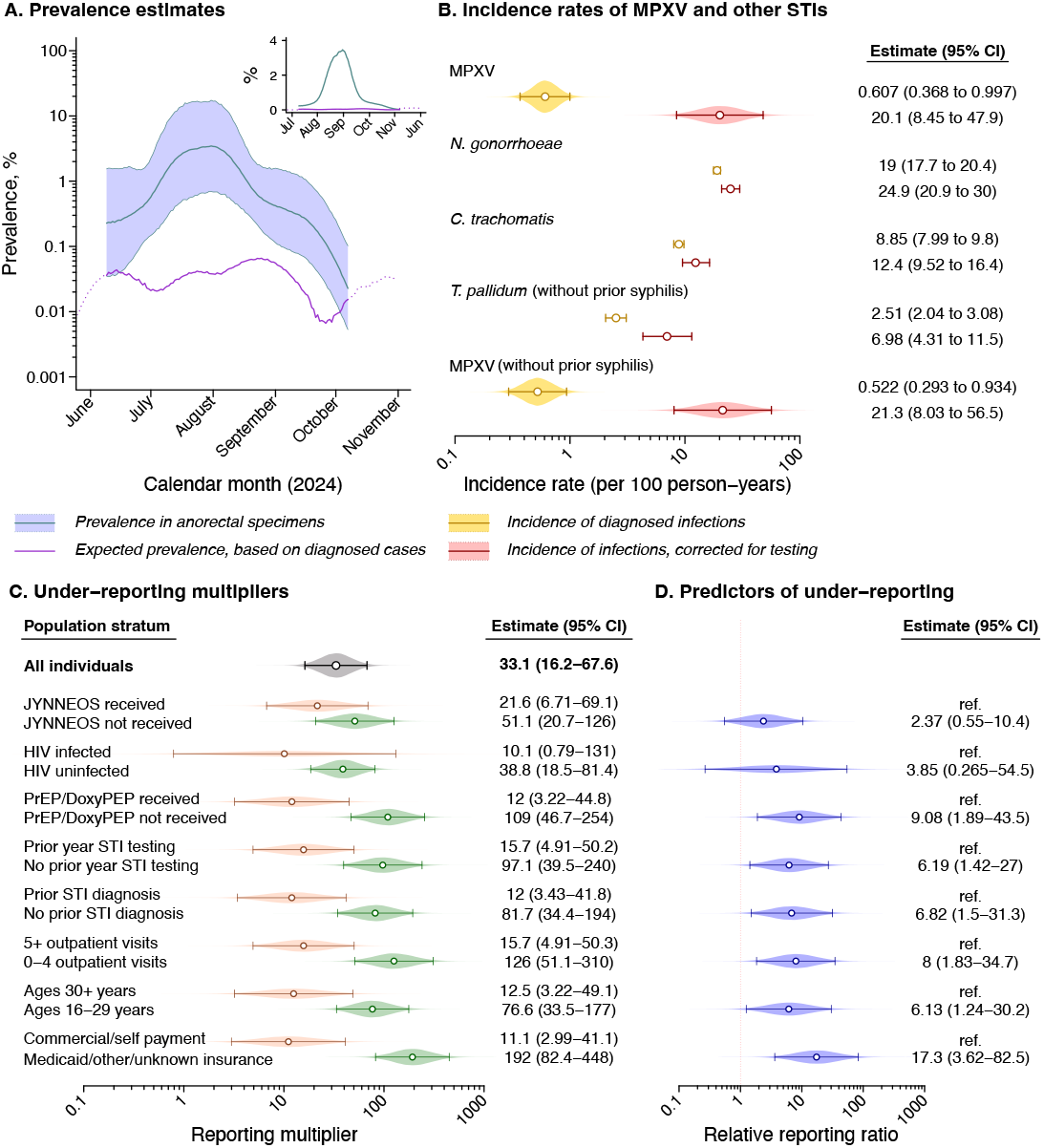
Prevalence and incidence of MPXV infection, and predictors of under-reporting. (**A**) We present estimates of expected prevalence of MPXV infection accounting only for shedding among cases receiving mpox diagnoses (purple line) alongside estimates of observed prevalence based on anorectal specimen testing (blue line, shaded area corresponding to 95% confidence intervals, presented as a moving 3-week average). Dotted lines convey prevalence estimates outside the period where a 3-week moving average could be computed for MPXV detection in anorectal specimens. We illustrate results on an absolute scale in the top-right inset panel. Estimates are weighted to adjust for differences in characteristics of individuals for whom anorectal specimens were collected or not collected during the study period. We detail c_*T*_ values for MPXV-specific and NVAR probes in **Table S3**. (**B**) Incidence rates of sexually-transmitted infections based on diagnosed cases (blue) and estimated rates of infection, corrected for testing effort (red). Rates are presented as infections per 100 person-years during the study period, within the weighted study cohort, for MPXV, *Neisseria gonorrhoeae, Chlamydia trachomatis*, and *Treponema pallidum*. Rates for *T. pallidum* are presented for individuals with no history of prior syphilis, due to use of serological diagnosis; rates for MPXV among individuals without prior syphilis diagnoses are presented below. (**C**) Estimates of the reporting multiplier, overall and for subgroups defined by demographic and clinical characteristics. We present alternative estimates based on an instantaneous prevalence comparison in **Table S4** along with results of sensitivity analyses excluding repeat positives or closely-spaced sampling intervals, and present results based on alternative parameterizations of the duration of shedding in **Table S5**. (**D**) Ratios comparing the estimated reporting multiplier across the distinct subgroups for which we present data.

No individuals testing positive for MPXV via anorectal specimen surveillance sought clinical care relating to mpox symptoms or received diagnostic testing during the study period, suggesting the infections we identified through rectal testing were asymptomatic or paucisymptomatic. In total, the 6 individuals testing positive for MPXV received 11 *N. gonorrhoeae*/*C. trachomatis* screening tests over the study period. One was receiving treatment for HIV infection, while 4 were receiving tenofovir-emtricitabine for HIV pre-exposure prophylaxis. The remaining individual not receiving antiviral treatment for HIV care or prevention was tested twice during the study period for sexually-transmitted infections (STIs), and received antibiotic treatment for both *N. gonorrhoeae* and *C. trachomatis* concurrently with his positive MPXV swab.

We next used a likelihood-based framework to estimate a reporting multiplier relating daily anorectal MPXV test positivity to expected rates of infection acquisition, shedding, and clearance derived from diagnosed cases.^1,17,22^ By this approach, we estimated that only 1 in 33 (16-68) MPXV infections received a laboratory-confirmed mpox diagnosis (**Figure 1C**).

Reporting multiplier estimates varied by individuals’ healthcare engagement and demographic characteristics; predictors of lower reporting included younger age, lack of HIV pre-exposure prophylaxis or doxycycline post-exposure prophylaxis, no prior-year STI testing or diagnoses, having <5 healthcare visits in the prior year, and receipt of health insurance through non-commercial sources (**Figure 1D**).

Sensitivity analyses excluding repeat positive results or closely spaced consecutive swabs (<30 days) yielded similar results, while an alternative, cross-sectional analysis comparing time-matched observed and expected MPXV prevalence estimated that 1 in 26 (12-47) infections was diagnosed (**Table S4**). Furthermore, point estimates from analyses using alternative data sources to parameterize the expected duration of anorectal MPXV shedding^23,24^ fell within the 95% confidence interval of our primary estimates (1 in 21 [11-43] to 1 in 52 [25-106]; **Table S5**). Our estimates also held when considering test specificity well below previously-published estimates,^25^ with risk of false positive results mitigated by our use of two distinct probes in qPCR assays (**Figure S3**).

Contrary to the low apparent incidence of mpox diagnoses, the estimated incidence rate of 20 (8.5-48) MPXV infections per 100 person-years within the KPSC study cohort was comparable to bacterial STIs for which MSM are recommended to receive routine testing. Estimated incidence rates of *N. gonorrhoeae* and *C. trachomatis* infection totaled 25 (21-30) and 12 (9.5-16) acquisitions per 100 person-years, respectively, over the same period (**Figure 1B**). Among persons without prior syphilis diagnoses, incidence of syphilis (*Treponema pallidum*) infections was substantially lower than incidence of MPXV infection (7.0 [4.3-11.5] versus 21 [8.0-57] acquisitions per 100 person-years), further highlighting the unexpected scale of MPXV transmission.

## Vaccination and MPXV infection

Among the study cohort, pre-exposure vaccination with JYNNEOS demonstrated 72% (9.4-91%) effectiveness against diagnosed mpox (78% for two doses, 55% for one dose; **Figure 2A**; **Table S6**), consistent with prior estimates.^26^ However, two different analysis frameworks suggested this effect was only partially mediated by protection against MPXV infection acquisition, with previously vaccinated individuals accounting for 5 of the 6 subclinical infections identified through anorectal specimen testing. Weighted incidence rates of MPXV infection were 51% (–92-95%) lower among JYNNEOS recipients than non-recipients (**Figure 2B**), and vaccination was associated with 53% (– 167-92%) effectiveness against anorectal MPXV infection within our surveillance study (59% for two doses, 41% for one dose; two-sided *p*>0.1). In turn, JYNNEOS recipients who acquired MPXV infection experienced 41% (–245-90%) lower risk of diagnosed mpox than unvaccinated individuals who acquired infection. Although imprecise due to low counts of observed subclinical infections, these estimates complement prior evidence that vaccination with JYNNEOS may reduce mpox severity without conferring sterilizing immunity against MPXV infection.^27–29^

**Figure 2:**
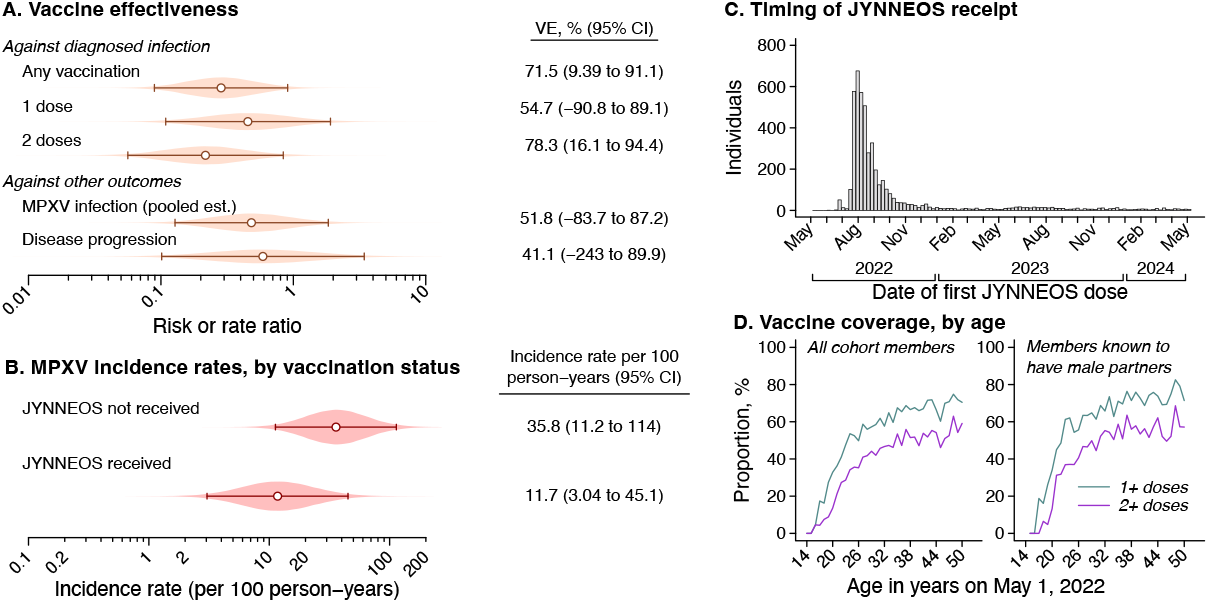
Testing-corrected incidence rates and vaccine effectiveness estimates. (**A**) Estimates of JYNNEOS vaccine effectiveness against diagnosed mpox, against MPXV infection, and against progression to diagnosed mpox, given MPXV infection. Effect estimates against diagnosed mpox are based on a case-control analysis comparing vaccination status among diagnosed mpox cases to individuals diagnosed with gonorrhea. Estimates of effectiveness against MPXV infection are pooled across analyses based on the incidence rate ratio of MPXV infection comparing vaccinated to unvaccinated individuals, and comparing prior vaccination among individuals testing positive or negative for MPXV infection by anorectal swabbing; estimates of effectiveness against disease progression are derived mathematically from estimated effectiveness against diagnosed mpox and against MPXV infection (**Table S7**). (**B**) Incidence rates of MPXV infection, corrected for testing effort, stratified by individuals’ history of JYNNEOS vaccination. (**C**) Dates of receipt of first JYNNEOS doses among vaccinated cohort members. (**D**) Coverage of ≥1 or ≥2 JYNNEOS doses among cohort members as of May 1, 2024, plotted against an axis indicating individuals’ age as of 1 May, 2022. Panels include all cohort members (left) as well as those with clinical notes indicating provider awareness of male sex partners (right).

Nearly all vaccinated individuals within the study population received JYNNEOS during acute phases of the mpox outbreak between July and October, 2022 (**Figure 2C**) amid expanded JYNNEOS delivery efforts.^30^ Low coverage among individuals aged <18 years in 2022 (12%) in comparison to individuals aged ≥18 years (60%) demonstrates ongoing growth of an undervaccinated cohort amid continued MPXV circulation (**Figure 2D**).

## Implications of subclinical infections for MPXV circulation

While these findings demonstrate that MPXV infections outnumber diagnosed cases, the contribution of subclinical or undiagnosed infections to MPXV transmission remains uncertain.^3,4,31^ To understand whether mpox epidemiologic observations are consistent with scenarios where individuals with undiagnosed infection contribute or do not contribute to onward spread, we next modeled MPXV transmission, accounting for dispersion in individuals’ number of secondary infections via a negative binomial offspring distribution (see **Methods**). Under a scenario in which undetected MPXV infections do not transmit infection,^3,32–34^ diagnosed mpox cases (representing 3.0% of total infections) would necessarily account for all onward spread. Such extreme concentration of transmission implies superspreading dynamics characterized by a very low dispersion parameter (*k*), which quantifies heterogeneity in infectiousness in epidemiological models.^35–37^

However, contrary to this scenario, phylogenetic analyses have estimated *k* values between 0.30-0.32 for MPXV transmission during the current clade IIb outbreak,^38–40^ similar to estimates for *N. gonorrhoeae* among MSM (*k*=0.26).^41^ Models excluding transmission by undetected infections—and minimizing dispersion by assuming uniform numbers of secondary infections among all diagnosed mpox cases—yielded maximal *k* estimates well below these previous estimates, ranging from 0.047 (0.021-0.092) to 0.082 (0.036-0.16) for scenarios with a reproduction number (*R*) between 0.7-3.5 (**Figure 3A**). Sensitivity analyses allowing transmission by 6.0% of the population—consistent with a higher reporting multiplier or 50% ascertainment fraction for individuals with clinical illness—likewise provided weak statistical support for observed patterns (2.3-11% probability for *k*≥0.3 with *R*<1, and <1% probability with *R*≥1).

**Figure 3:**
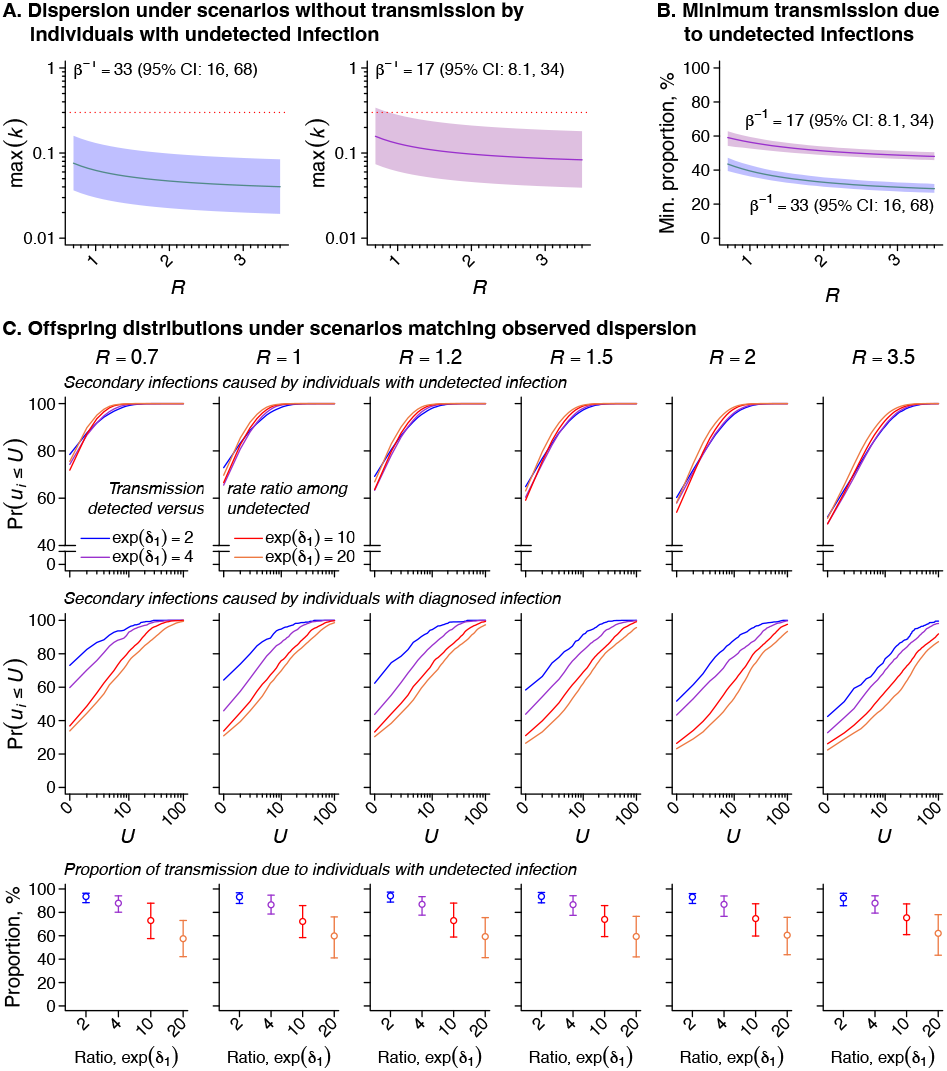
Modeling the contribution of individuals with diagnosed mpox and undetected MPXV infection to transmission. (**A**) Maximum values of the dispersion parameter, *k*, obtained under scenarios where only individuals with detected infections transmit MPXV. Modeled scenarios minimize dispersion in the offspring distribution by assuming uniform numbers of secondary infections for the proportion (1 ™ *β*) of individuals with diagnosed mpox, and no secondary infections for the proportion *β* of individuals with undetected MPXV infection. The reproduction number *R* denotes the mean number of secondary infections caused by each infected individual. We consider scenarios with reporting multipliers of *β*^−1^ = 33 (per our primary analysis) and *β*^−1^ = 17 (as a sensitivity analysis). Shaded areas delineate 95% confidence intervals around our estimates, and dotted red lines correspond to *k* = 0.3, as estimated in previous studies.^38–40^ (**B**) Minimum proportion of transmission attributable to undetected infections, under scenarios with *k* = 0.3 and deterministic assignment of transmission links to index cases with diagnosed mpox. We present estimates with reporting multipliers of *β*^−1^ = 33 (per our primary analysis) and *β*^−1^ = 17, as a sensitivity analysis. (**C**) Offspring distribution characteristics under scenarios with differing transmission dynamics. Top and middle rows present cumulative distribution functions for the number of secondary infections (*U*) caused by individuals with undetected MPXV infection and diagnosed mpox, respectively, varying the rate ratio of transmission by individuals with diagnosed mpox relative to those with undetected MPXV infection, exp (*δ*_1_), from 2 to 20. Bottom row panels indicate the total proportion of transmission attributable to individuals with undetected infection for each value of *δ*_1_. Panels are grouped into columns according to *R* values employed for each analysis (*R=*0.7, 1.0, 1.2, 1.5, 2.0, 3.5).

We therefore quantified the contribution of undetected infections to transmission under more realistic scenarios with MPXV dispersion characteristics mirroring prior observations (*k*=0.3). Maximizing the proportion of transmission attributable to diagnosed cases via a deterministic framework, we estimated that individuals with undetected infection must account for a minimum of 31% (29-34%) to 44% (40-47%) of all transmission events (**Figure 3B**). This estimated attributable fraction exceeded half of all transmission under less-restrictive modeling assumptions allowing differences in infectiousness among individuals with diagnosed or undiagnosed infections (**Figure 3C**). Assuming individuals with diagnosed infection are 20-fold more likely to transmit MPXV compared to undetected cases, the proportion of transmission attributable to undetected infections was 61% (43-78%). Assuming weaker differences in infectiousness (only a 2-fold increase for diagnosed cases relative to undetected infections), the contribution of undetected infections rose to 94% (88–97%). Thus, to explain dispersion patterns evident in MPXV phylogenies, undetected infections must play a meaningful role in sustaining viral transmission.

Current guidance defines elimination of human-to-human transmission as the absence of new, locally-acquired cases (without travel history or zoonotic exposure) for ≥3 months in settings with adequate capacity to conduct laboratory testing for individuals meeting clinical case criteria.^8^ However, at our estimated reporting multiplier, the probability of observing 0 cases would exceed 50% under scenarios where up to 23 (11-47) infections had occurred, and would exceed 5% with as many as 98 (48-202) true infections (**Figure 4A**). In modeling analyses considering our estimated reporting fraction of 1 in 33 infections, presence of 0 diagnosed cases over a 3-month span was unlikely to indicate absence of ongoing transmission under most scenarios with *R*≥1 (**Figure 4B**). With *R*=1.1, the probability of containment after 3 months without detected cases was 25%, 13%, 4.5%, 1.2%, and 0.14% in scenarios where this lapse in detections was preceded by outbreaks involving 5, 10, 25, 50, and 100 or more notified cases, respectively. The same probabilities were <10% for scenarios with *R*≥1.2, and <1% for scenarios with *R*≥1.5. Because undetected MPXV circulation may persist over extended periods without clinically-detected cases, expanded surveillance efforts including asymptomatic testing in high-risk populations may be necessary to monitor transmission.

**Figure 4:**
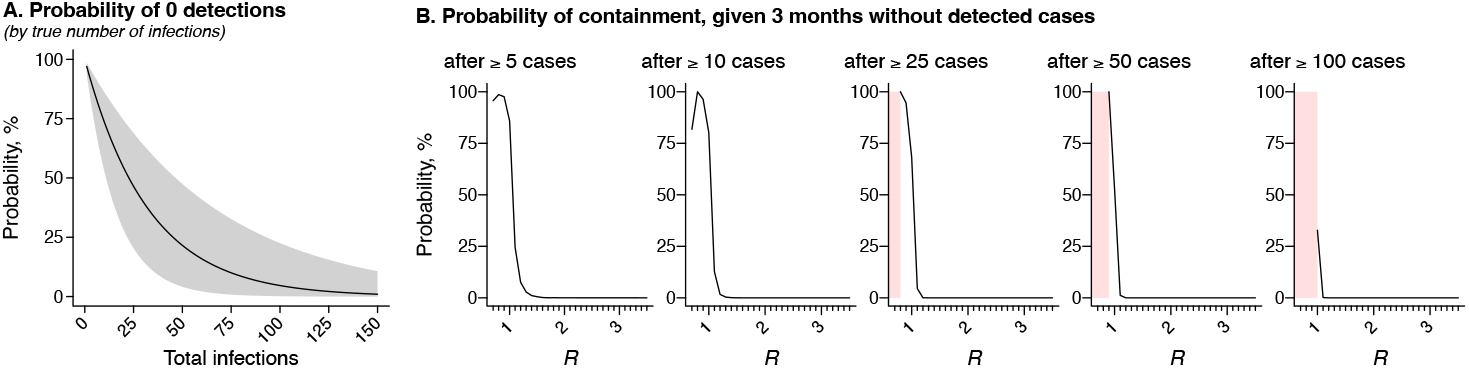
Elimination thresholds for mpox. (**A**) Probability of observing 0 diagnosed mpox cases, based on true number of infections, assuming 3.0% (1.5-6.2%) of infections receive diagnoses. The shaded area delineates 95% confidence bounds around point estimates (line). We generate estimates assuming the number of diagnosed cases is a binomial random variable. (**B**) Probability of containment, defined as the cessation of ongoing transmission 90 days after the last notified case. Panels illustrate the estimated probability of containment, generated via simulation of transmission chains with a negative-binomial offspring distribution and varying values of *R*, under scenarios where ≥5, ≥10, ≥25, ≥50, and ≥100 notified cases occurred prior to the last notified case. Shaded pink areas delineate ranges of *R* where all simulated transmission chains were depleted before the minimum number of notified cases occurred.

## Under-reporting is geographically widespread and confirmed by phylogenetic analyses

To understand the external generalizability of our results, we next compared evidence of under-reporting within the KPSC cohort to data from other settings. Numerous surveillance studies have identified undetected MPXV infections based on anorectal shedding or serological evidence of prior infection, although implications of these findings for local mpox reporting fractions remain unknown. We undertook a systematic review of epidemiological studies conducting molecular or antibody-based testing for MPXV infection among MSM, and compared findings from 10 studies meeting eligibility criteria^16,18,33,34,42–47^ to mpox case notifications from the same settings (**Tables S7**-**S9**; **Figure S4**).

In studies testing for MPXV shedding via anorectal specimens^16,18,34,42–44^, observed prevalence exceeded expected prevalence based on reported cases by factors of 51-525 (**Figure 5A**). Adjusting for potential higher-risk behavior in tested populations (**Figure S4**), estimated reporting multipliers ranged from 1 in 36-377. Serological studies revealed a lower, but still substantial, extent of under-reporting (1 in 7.0-69 infections, unadjusted; 1 in 5.0-50, adjusted; **Figure 5B**).^33,34,45–47^ In the single study employing both molecular and serological testing,^34^ reporting multipliers estimated from serological testing revealed 4.0-fold (0.86-15) greater reporting completeness than estimates based on molecular testing results, suggesting differences were driven by lower sensitivity of serological testing.

**Figure 5:**
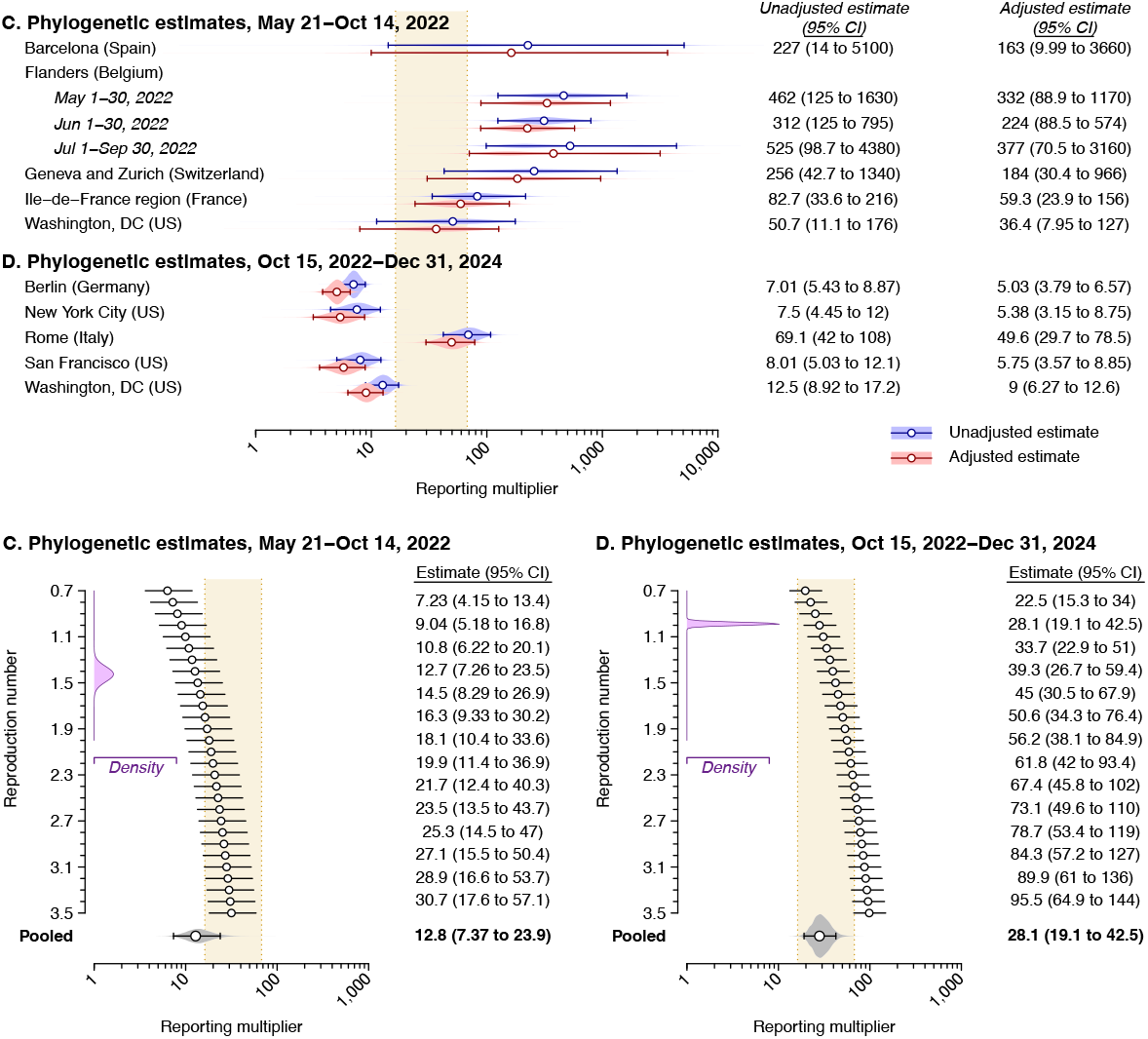
Reporting multiplier estimates based on external surveillance studies and phylogenetic analyses. (**A**) Estimated reporting multipliers comparing MPXV infection prevalence observed in anorectal specimen testing to expected prevalence based only on diagnosed cases in each study setting. We detail the included studies and data providing the basis for estimates in **Table S7** and **Table S8**. Risk-adjusted estimates account for expected differences in MPXV detection in samples recruited in STI clinic settings as compared to the general MSM population (**Figure S5**). The tan shaded area delineates the 95% confidence interval around our primary estimate within the KPSC study population (33; 95% confidence interval [CI]: 16-68) as a basis for comparison. (**B**) Estimated reporting multipliers comparing anti-MPXV IgG seroprevalence to expected seroprevalence based on cumulative diagnosed cases in the study setting. We present pooled estimates of reporting multipliers across studies, with and without risk adjustment, in **Table S9**. (**C**) Reporting multipliers based on a comparison of diagnosed cases to the estimated population size of MPXV infections in Los Angeles County from phylogenetic analyses of sequenced cases, for the period from May 21 to October 14, 2022. Estimates are conditioned on the reproduction number, *R*. We illustrate the probability density of mean daily *R* values throughout the analysis period in purple (top left inset panel). Pooled estimates presented in bold at the bottom of the figure are weighted according to the period-specific probability density of *R*. We present alternative estimates assuming lower or higher values of the dispersion parameter *k* in **Figure S7**. (**D**) Estimated reporting multipliers comparing diagnosed cases to the estimated population size of MPXV infections in Los Angeles County, based on phylogenetic analyses, for the period from October 15, 2022 to December 31, 2024.

To further validate our findings using a distinct analytical framework and data source, we next estimated reporting multipliers via phylogenetic reconstruction of MPXV transmission within Los Angeles County. Leveraging data from 497 clade IIb MPXV genomes sampled locally alongside 7,140 publicly available global sequences collected between May 2022 and December 2024, we fit a birth-death skyline model^48^ to local outbreak clusters using Markov chain Monte Carlo in BEAST2, as described previously,^38,39,49^ and used estimates of the sampling proportion to compute reporting multipliers.^50^ Validating this framework for estimating the sampling proportion in the presence of transmission heterogeneity (**Figure S6**), we estimated that MPXV infections outnumbered reported cases by a factor of 28 (19-43) from October 15, 2022 to December 31, 2024, closely resembling our primary estimate of 33 (16-68) based on anorectal specimen testing. We also estimated that MPXV infections exceeded reported cases by a factor of 13 (7.4-24) during the preceding emergency phase of the outbreak (**Figure 5C-D**; **Figure S7**). This 49% (23-96%) reduction in the proportion of MPXV infections receiving diagnoses after October 15, 2022 aligns with the rapid rollout of JYNNEOS for at-risk MSM, further suggesting a role of vaccination in attenuating disease severity: 76% of Los Angeles County recipients of JYNNEOS,^51^ and half of the full KPSC study cohort, were vaccinated by October 15, 2022.

## Discussion

Our results demonstrate frequent undetected clade IIb MPXV infections among MSM, with infections outnumbering diagnosed cases by a 33-fold margin within a cohort of Los Angeles MSM during summer, 2024. Estimated reporting multipliers spanned 1 in 21-52 in analyses using alternative parameterizations of individuals’ duration of infection, while independent datasets derived from molecular surveillance, serological studies, and phylogenetic analyses supported this finding of extensive under-reporting. Most subclinical infections occurred among individuals who previously received JYNNEOS, in agreement with prior evidence that vaccination may reduce disease severity^27–29^ rather than providing sterilizing immunity against MPXV. The low estimated reporting fraction further implies that undetected infections contribute substantially to transmission, contradicting current guidance that suggests undetected infections are uncommon^4,31,32,32^ and emphasizes symptom-driven transmission.^3^ Our outcomes suggest that currently-endorsed elimination thresholds based on the absence of reports of locally-acquired cases for ≥3 months may provide a poor proxy for resolution of outbreaks. Ongoing cryptic circulation of MPXV may undermine the effectiveness of currently-endorsed public health strategies focused on identifying cases with clinical illness and tracing their contacts,^8–11^ and underscores the importance of renewed vaccination efforts to protect populations at high risk of exposure.

To our knowledge, no prior studies have directly estimated the proportion of MPXV infections that are clinically diagnosed based on results of symptomatic and asymptomatic testing. A study prospectively monitoring contacts of mpox cases for MPXV infection reported symptoms among 46% of individuals who tested positive,^17^ although intensive monitoring likely enhanced case ascertainment among individuals with mild clinical presentations in this study. Across multiple molecular screening studies included in our meta-analysis,^16,18,34,42–44^ 22% (11/49) of MPXV-positive participants who were followed up reported symptoms or received clinical diagnoses (**Table S10**), while a seroprevalence study from 2022 reported that only 13% of individuals with evidence of prior infection recalled recent rash or lesions.^45^ In a 2022 study of US MSM, 18% and 7% of participants who experienced rash with and without fever during the outbreak peak (June-August), respectively, were tested for mpox,^52^ suggesting case ascertainment may be suboptimal even among individuals who experience symptoms.

Our findings augment a growing body of observations compatible with MPXV transmission by individuals with inapparent infection. Consistent with our finding that undetected infections account for at least 31-44%, and likely 61-94%, of transmission, 67-85% of mpox cases in previous studies have reported no known exposure to individuals with mpox or mpox-like symptoms.^2,12–15^ Furthermore, 28-77% of transmission events in studies of linked mpox cases involved pre-symptomatic exposures, confirming that contact with lesions is not requisite for spread.^2,19,21^ Shedding of replication-competent MPXV occurs before symptoms onset and during asymptomatic infection in oral, anal, and genital mucosa, providing a plausible mechanism of transmission besides skin-to-lesion contact.^16–18^ Last, infection risk is strongly associated with condomless anal or vaginal sex,^2,47,53^ which may be incompatible with the severe pain cases report while experiencing lesions.^54^

Monitoring MPXV shedding and incident disease in a single cohort, with detailed demographic and healthcare utilization data, enabled direct comparison of MPXV shedding prevalence to mpox diagnoses. Reporting multipliers based on estimates from molecular and serological surveillance studies, as well as phylogenetic analyses, validated our conclusions, and our results were robust to varied parameterizations of MPXV natural history and test specificity. However, our study has limitations. For at least two reasons, our estimates likely represent a lower-bound for the extent of under-reporting of MPXV infections within the KSPC study cohort. First, although anorectal swabs are optimal for identifying subclinical MPXV infections,^17,55^ studies using multiple specimen sources have identified further infections based on oropharyngeal swabs alone.^18,53^ Second, available evidence suggests that greater symptom severity corresponds to longer duration of anorectal MPXV shedding.^24^ While direct comparisons of the duration of shedding among individuals with symptomatic and asymptomatic infections are not available, our analyses may underestimate the incidence of subclinical infections if underlying parameters derived from clinically-diagnosed cases overestimate the duration of asymptomatic infection. Separately, with 6 infections identified by anorectal specimen testing, our assessments of vaccine effectiveness against infection were underpowered; nonetheless, consistency of our point estimates under two analysis frameworks supports their validity. We also did not assess duration of vaccine protection, as most JYNNEOS recipients were vaccinated within a short period during 2022. Our analyses of prior molecular and serosurveillance studies lacked individual-level metadata, and relied on uncertain estimates of MSM population denominators for each setting, likely contributing to wide variation in estimates. Differences in estimates based on molecular and serological testing may indicate greater sensitivity of PCR testing. This could arise if weaker seroresponse among individuals with subclinical MPXV leads to false-negative results of serological testing, especially for assays calibrated with sera from symptomatic cases. Last, differential sequencing efforts globally, and non-random sequencing of transmission clusters in Los Angeles County, could impact our ability to infer MPXV introductions and estimate transmission-dynamic parameters within phylogenetic analyses.

Collectively, our results challenge prevailing assumptions underlying symptom-based mpox surveillance and support reconsideration of control strategies in light of widespread and clinically inapparent MPXV circulation. Surveillance efforts relying on lesion-based clinical testing do not accurately convey individuals’ likelihood of exposure to MPXV among sexual contacts, and declining case notifications should not be interpreted as evidence of successful containment.^5^ Public health strategies reliant on symptom-based testing, isolation, and contact tracing are unlikely to be effective in curtailing MPXV transmission, and the current WHO-endorsed elimination threshold^7^ may not indicate true cessation of spreading. After relaxation of enhanced efforts to distribute JYNNEOS during emergency phases of the mpox response, young MSM, in particular, have low vaccine coverage and may be at risk of severe mpox amid ongoing exposure to MPXV circulation. Long-term control strategies ensuring continued access to JYNNEOS and reliable monitoring of infection prevalence are needed to manage MPXV as an endemic STI among MSM.

## Methods

### Study cohort

Eligible individuals were males born between January 1, 1972 and May 1, 2008 who had been members of KPSC health plans continuously since at least 1 May, 2023 (allowing enrollment gaps <45 days), ensuring capture of prior-year healthcare utilization and recent STI history. Our age-based restrictions limited the population to individuals born after cessation of smallpox vaccination for the US general public, improving comparability of our estimates to those of other prospective testing studies^33,47^ and constraining the sample to age groups experiencing the greatest risk of mpox and other STIs.^13^ We further restricted eligibility to individuals who: received ≥1 test anorectal *N. gonorrhoeae*/*C. trachomatis* test after 1 May, 2023, as such testing is indicated only for MSM, and who over the same period had any appointment or received any laboratory test, prescription fill, immunization, or other care at the West Los Angeles Medical Center or Los Angeles Medical Center. As these were the KPSC facilities diagnosing the greatest number of mpox cases since 2022, individuals receiving care from these facilities were expected to have the greatest likelihood of mpox diagnosis in the event of clinical illness.

### Testing

Screening for STIs is recommended every 3-6 months for MSM receiving HIV pre-exposure prophylaxis and those living with HIV who continue to have new sex partners, and for MSM receiving doxycycline post-exposure prophylaxis.^56,57^ Between May 29 and November 13, 2024, we obtained a total of 1,190 remnant anorectal specimens submitted for *N. gonorrhoeae*/*C. trachomatis* screening among 1,054 of the 7,930 study cohort members during visits to the West Los Angeles or Los Angeles Medical Center facilities. Samples were maintained at –20ºC and tested at the Southern California Permanente Medical Group, Regional Reference Laboratories, Chino Hills, using a clinically-validated multiplex real-time PCR that detects MPXV (clades I/II) and NVAR, as previously described.^25,58^ We defined confirmed MPXV infections as those with positive results for both MPXV (c_*T*_≤36.9) and NVAR (c_*T*_≤37.6), consistent with previously-established lower limits of detection; no specimens within the study sample had inconsistent results across probes. We list primer and probe sequences for both tests in **Table S11**. We included an internal PCR control, i.e., a non-target nucleic acid sequence, in each reaction, and tested specimens in batches of up to 94, including positive and negative external controls within each batch.

### Sample weighting and prevalence estimation

To address differences in characteristics of individuals from whom we received or did not receive a specimen for MPXV testing during the study period, we adjusted estimates of mpox incidence and MPXV infection prevalence within the KPSC study cohort via stabilized inverse propensity weighting.^59^ We computed stabilized inverse propensity-of-testing weights (*ω*_*i*_) using a logistic regression model defining receipt of testing as the outcome; covariates included individuals’ age group, race/ethnicity, health insurance payment source, neighborhood deprivation index (a community-level proxy for individual socioeconomic status), prior-year healthcare utilization across outpatient settings and history of emergency department presentation or inpatient admission, prior-year STI testing, receipt of HIV pre-exposure prophylaxis, receipt of doxycycline post-exposure prophylaxis, receipt of JYNNEOS, prior diagnoses of HIV, syphilis, gonorrhea, chlamydia, mpox, or other STIs, and prior diagnosis of alcohol or drug abuse.

### Time-to-event analysis for under-reporting

To generate expected infection prevalence estimates based on diagnosed mpox cases, we sampled dates of MPXV exposure (*τ*_*E*_), shedding onset (*τ*_*S*_), symptoms onset (*τ*_Y_) and shedding cessation (*τ*_*C*_) for cases based on their dates of testing (*τ*_*L*_). We estimated daily infection prevalence within the study cohort (*π*(*t*)) by dividing the sum of individuals expected to shed MPXV in anorectal specimens by the total enrolled population, applying weights as defined above, via

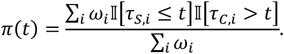

We abstracted data on duration of several stages within the natural history of MPXV infection to parameterize distributions for sampling event times *τ*_*S*_ and *τ*_*C*_ from observations *τ*_*L*_ (**Figure S2**). From a study of reported cases followed prospectively from the date of diagnostic testing with repeated sampling,^22^ we estimated time from symptoms onset to cessation of anorectal viral DNA detection detectable by PCR (*τ*_*C*_ − *τ*_*Y*_). From a case series of US mpox patients,^1^ we estimated times from symptoms onset to testing (*τ*_*L*_ − *τ*_*Y*_). Last, from a study that prospectively followed high-risk contacts of confirmed mpox cases,^17^ we estimated times from exposure to onset of anorectal viral DNA detection detectable by PCR (*τ*_*S*_ − *τ*_*E*_), as well as times from exposure to symptoms onset (*τ*_*Y*_ − *τ*_*E*_). We assumed Gamma-distributed times to event and used maximum likelihood estimation to fit distribution parameters to reported data; where authors presented information on distribution quantiles only, without accompanying individual-level data, we fit Gamma distribution parameters minimizing summed squared errors between expected and reported distribution quantiles.

Our primary analysis related daily weighted, observed prevalence of infection within the anorectal swabbing study to expected infections based on reported cases in a likelihood-based framework. We defined the probability of experiencing onset of anorectal shedding on day *k* as

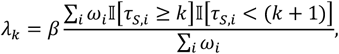

where *β* represented a multiplier relating the true number of individuals experiencing onset of shedding on day *k* to the expected number of individuals experiencing onset of shedding on day *k* based on diagnosed cases alone. Conditioned on experiencing shedding onset on day *k<t*, we defined the probability an individual *j* would be observed to shed on day *t* as Pr[*Z*_*j*_(*t*) = 1|*k*, = Pr[(*τ*_*C*_ − *τ*_*S*_) > *k*]. The probability of not shedding on day *t* was thus the probability of not initiating shedding by day *t*, or experiencing shedding onset and cessation before day *t*:

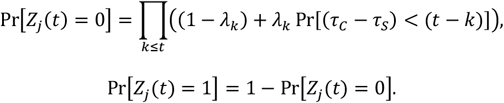

The weighted likelihood all *Z*_*j*_(*t*) observations was thus

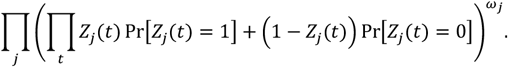

Extending this framework for analyses comparing under-reporting between participant strata distinguished by a binary characteristic *X* ∈ {0,1}, as presented in **Figure 1**, we defined

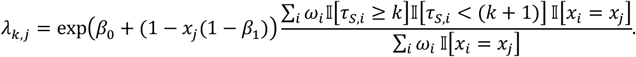

We estimated the reporting multiplier *β* (or *β*_0_ and *β*_1_, for stratified analyses) by maximum likelihood, obtaining the covariance via the inverse of the Hessian matrix.

### Alternative parameterizations of the duration of shedding

We identified two additional studies providing an indirect basis for quantifying the duration of anorectal MPXV shedding following symptoms onset. The first^23^ presented viral loads from 162 anorectal specimens collected 1-19 (median: 19; interquartile range: 6-12) after symptoms onset. We fit a regression model relating log-transformed viral load measurements to days from symptoms onset (weighting individuals by HIV status to resemble HIV status among those testing positive for MPXV in the KPSC study cohort). We confirmed via comparison of Bayesian information criterion values that analyses defining time as a continuous variable outperformed analyses applying log, quadratic, and cubic transformations of time. We then generated 10,000 draws from the prediction interval of individuals’ viral load measurements on days 1-100 after symptoms onset to estimate the probability that values would be above the lower limit of detection each day (corresponding to c_*T*_=37 in the primary study). We used these probabilities to define the cumulative distribution function for time to cessation of shedding, and computed first differences in the cumulative distribution function to obtain a corresponding probability mass function for days to cessation of shedding. The resulting estimate (median: 27 days; interquartile range: 17-56 days) exceeded the estimate used in primary analyses (median: 16 days; interquartile range: 12-21 days; **Figure S2**), yielding a reporting multiplier of 1 in 21 (11-43) infections (**Table S5**).

The second study^24^ presented results from PCR testing for anorectal MPXV shedding in 18 men diagnosed with mpox, among whom all received an initial test within 1-7 days after symptoms onset and 9 were retested within 21-32 days after symptoms onset. We fit a gamma distribution for time to cessation of shedding maximizing the likelihood of the observed numbers of individuals continuing to test positive by the time each sample was collected (median: 9.7 days; interquartile range: 6.7-14 days), yielding a reporting multiplier of 1 in 52 (25-106) infections (**Table S5**).

### Comparison of observed to expected infection prevalence

We conducted an alternative analysis comparing observed to expected prevalence throughout the study period. We sampled values of the mean expected prevalence throughout the study period, 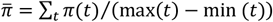, via draws of *τ*_*S*_ and *τ*_*C*_ for each case, and sampled values of the observed prevalence within the anorectal specimen study as *p* ~ Beta (∑_*j*_∑_*t*_ *ω*_*j*_*z*_*j*_(*t*), ∑_*j*_∑_*t*_ *ω*_*j*_ (1 − *z*_*j*_(*t*))). We defined our alternative estimate for the reporting multiplier 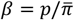, and sampled from *β* via 100,000 independent draws from *p* and 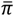.

### Test specificity considerations

We conducted several analyses aiming to explore potential implications of false-positive detections for our results. Previous real-world validation of the study assay^25^ observed *x*=0 positive results among *n=*64 true negative specimens; parameterizing a corresponding negative binomial distribution under differing specificity values (*p*) ranging from 0.1% to 15% for a single test, the maximum likelihood value for single-test specificity was 100%, and single-test specificity was expected to take values ≥99.9%, ≥98.8%, and ≥94.8% with likelihood of 90%, 50%, and 5%, respectively. We quantified the positive predictive value (PPV) of a dual-positive result for MPXV and NVAR as

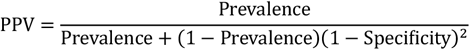

under scenarios with true prevalence equal to 0.91% (as estimated in the study) or 0.455% (corresponding to a scenario where half of all detections were false positives). We repeated our primary analyses multiplying infection prevalence by the resulting dual-positive PPV to generate corrected estimates accounting for varying test specificity (**Figure S3**).

We also used the distribution of quantitative c_*T*_ values for the MPXV-specific and NVAR probes to assess potential false-positive signals. Whereas c_*T*_ values were expected to be correlated if measuring true MPXV-associated nucleic acids, spurious signals leading to false positive results were expected to have no association across probes. To simulate the expected difference in MPXV-specific and NVAR c_*T*_ values under a scenario of independence (due to false positive results), we took 7 draws each from the distributions of observed c_*T*_ values for each probe. We computed mean differences in c_*T*_ between the two probes, and measured the probability of a mean difference as extreme as that observed in our sample as

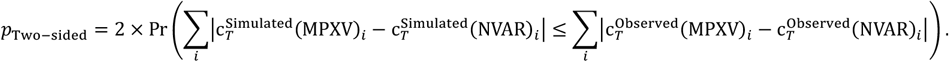

### Incidence rate estimation

We estimated incidence rates of MPXV infection within the reweighted sample, adjusted for reporting, as *β*∑_*i*_ *ω*_*i*_*y*_*i*_/∑_*i*_ *ω*_*i*_ min(*τ*_*Y,i*_, *τ*_*F,i*_) where *τ*_*F,i*_ was the earliest of the date of death, disenrollment, or study termination for individual *i*.

To contextualize our estimates of the incidence rate of MPXV infection, we also estimated incidence rates for infection with *N. gonorrhoeae, C. trachomatis*, and *T. pallidum*. We defined *N. gonorrhoeae* and *C. trachomatis* infections as any positive molecular test result separated by ≥30 days from any prior positive test result or diagnosis; we defined *T. pallidum* infection based on positive results for *Treponema*-specific IgG or IgM. To correct for potential under-ascertainment of these other STIs, we fit Poisson regression models relating individuals’ rates of infection during the study period to their screening frequency in the prior year. Models defined counts of unique diagnoses (separated by ≥30 days from a previous diagnosis) with each infection during the study period as the outcome variable and included log-transformed person-time at risk as an offset term. The primary exposure of interest was the prior-year frequency of testing (between May 1, 2023 and April 30, 2024). For *N. gonorrhoeae* and *C. trachomatis, w*e defined receipt of ≥4 tests as the “best-case” referent category based on guidelines that MSM should be screened up to every 3 months for continuation of HIV PrEP and doxycycline post-exposure prophylaxis;^56,57^ for *T. pallidum*, we defined receipt of ≥2 tests as the “best-case” referent category due to the lower rate of infection and lack of evidence for increased incidence of syphilis diagnoses with testing at shorter intervals in the study population. To ensure measured testing behavior reflected engagement with screening rather than test-seeking for symptomatic illness or known exposures, models adjusted for individuals’ receipt of any gonorrhea diagnosis or chlamydia diagnosis in the prior year, as well as age group, insurance source, receipt of HIV PrEP and doxycycline post-exposure prophylaxis, receipt of JYNNEOS, HIV infection and prior syphilis diagnosis. As *T. pallidum* infection is diagnosed by serology, we limited analyses for this infection to individuals with no prior history of syphilis diagnoses or positive test results.

Regression parameters *α*_1_ and *α*_2_ compared rates of STI diagnoses associated with prior-year receipt of 0-1 tests versus ≥4 tests and 2-3 tests versus ≥4 tests, respectively, and thus represented reporting ratios for STI compared to a scenario where individuals’ testing frequency met CDC recommendations. We defined the correction factor relating true rates of infection to rates of diagnoses as

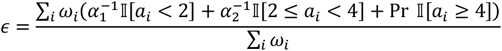

and multiplied incidence of rates of STI diagnoses by *ϵ* to obtain incidence rates of infection. The corresponding equation for corrected incidence rates of *T. pallidum* infection was

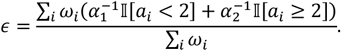

### Vaccine effectiveness estimation: overview

We aimed to estimate three classes of vaccine direct effects comparing counterfactual risk of mpox-related clinical outcomes among individuals who received JYNNEOS versus those who did not receive JYNNEOS. Following Halloran et al.,^60^ we considered that vaccination could protect against diagnosed mpox by: reducing individuals’ susceptibility to acquiring MPXV infection, given exposure, by a factor VE_*S*_ = (1 − *θ*_*S*_) × 100%; and by reducing individuals’ risk of progression to diagnosed mpox, given MPXV infection, by a factor VE_*P*_ = (1 − *θ*_*P*_) × 100%. The reduction in risk of diagnosed mpox, given vaccination, was thus VE_*D*_ = (1 − *θ*_*S*_*θ*_*P*_) × 100%.

We used a case-control design to estimate VE_*D*_, and confirmed results of this primary analysis using a negative control-adjusted prospective cohort framework. We also estimated VE_*S*_ using two analysis frameworks, based on both case-control and prospective-cohort designs. We used the results of these analyses to estimate VE_*P*_, defining

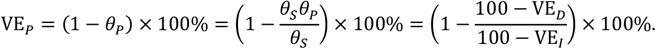

Our analyses considered pre-exposure JYNNEOS vaccination only, as no post-exposure prophylaxis with JYNNEOS occurred among cohort members during the study period. All previously-administered JYNNEOS doses were considered pre-exposure doses with respect to follow-up for mpox during the study period.

### Estimation of vaccine effectiveness against diagnosed mpox

Our primary analysis estimating VE_*D*_ used a case-control framework comparing adjusted odds of prior vaccination among cases diagnosed with laboratory-confirmed mpox to controls diagnosed with laboratory-confirmed gonorrhea during the study period. This strategy was anticipated to mitigate confounding based on the expectation that receipt of JYNNEOS could be associated with individuals’ risk of STI exposure as well as their engagement with sexual health services and likelihood of being diagnosed, if infected;^61,62^ whereas JYNNEOS would not be expected to alter individuals’ risk of gonorrhea, gonorrhea cases were expected to resemble mpox cases in sexual risk characteristics and healthcare-seeking behavior. We defined the product *θ*_*S*_*θ*_*P*_ as the adjusted odds ratio of prior JYNNEOS vaccination (receipt of any doses, 1 dose, or ≥2 doses) among mpox cases relative to controls diagnosed with gonorrhea, and estimated this term via conditional logistic regression. We defined matching strata on individuals’ HIV infection status and (among HIV-negative individuals) receipt or non-receipt HIV PrEP; receipt of doxycycline post-exposure prophylaxis; and history of any syphilis diagnosis. Models further controlled for individuals’ age group, receipt of *N. gonorrhoeae*/*C. trachomatis* testing in the prior year, and commercial or non-commercial insurance source (expected to proxy socioeconomic status) via covariate adjustment.

We conducted a sensitivity analysis leveraging the prospective design of the study and estimating VE_*D*_ via a negative control-corrected incidence ratio, following a previously-described framework^63^ recently validated in a randomized controlled trial setting.^64^ This analysis used a two-stage approach; we first estimated adjusted incidence rate ratios (IRRs) comparing incidence of mpox diagnoses and gonorrhea diagnoses among JYNNEOS recipients (any receipt, 1 dose receipt, or ≥2 dose receipt) versus non-recipients. Considering that any apparent association of JYNNEOS vaccination with gonorrhea incidence would signify bias due to differential sexual risk and healthcare-seeking behaviors among vaccine recipients and non-recipients, we used the estimated IRR for gonorrhea to adjust the causal effect estimate for the IRR of mpox associated with JYNNEOS vaccination, such that

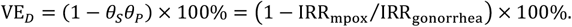

We estimated each IRR via Poisson regression models, defining mpox or gonorrhea diagnoses as the outcome variables and including log person-time at risk as an offset, and adjusted for all risk factors included in the primary analysis models (HIV infection, receipt of HIV PrEP, receipt of doxycycline post-exposure prophylaxis, prior syphilis diagnosis, age group, prior-year gonorrhea testing, and commercial insurance source) as covariates.

### Estimation of vaccine effectiveness against MPXV infection

Our first analysis estimating VE_*S*_ defined *θ*_*S*_ as the adjust odds ratio of prior JYNNEOS receipt (any doses, one dose, or two doses) or non-receipt comparing individuals within the anorectal specimen testing study who received positive MPXV results to those receiving negative results. We considered the outcome of positive results from asymptomatic rectal specimen based on the assumption that infections captured within the testing study represented the source population from which diagnosed mpox cases would be drawn; although none of the 6 individuals receiving positive results in our study ultimately progressed to diagnosed illness, such progression has been identified in other studies employing similar designs (**Table S10**). We fit *θ*_*S*_ using conditional logistic regression models defining positive or negative test results from each specimen as the outcome, defining matching strata for individuals’ HIV infection status and (among HIV-negative individuals) receipt or non-receipt HIV PrEP, receipt of doxycycline post-exposure prophylaxis, and history of any syphilis diagnosis, consistent with our analyses estimating VE_*D*_. We included data from all specimens, and used the sandwich estimator to adjust variance estimates for repeat sampling of individuals.

We also estimated VE_*S*_ via the adjusted IRR of MPXV infection comparing previously-vaccinated individuals to unvaccinated individuals. These analyses applied reporting multipliers (*β* parameters, as described above) specific to the vaccinated and unvaccinated populations in defining MPXV incidence rates for the comparator populations.

### Examining the role of undetected infections in transmission: overview

We undertook two analyses aiming to clarify the potential contribution of individuals with undetected MPXV infections to transmission. The first quantified the minimum degree of dispersion (maximum value of *k*) corresponding to a scenario where individuals with undetected infection make no contribution to transmission, and compared this value to previously-reported estimates of *k* for MPXV based on prior phylogenetic studies.^38–40^ The second analysis estimated the proportion of infections caused by individuals with undetected infection under scenarios with *k* = 0.3, consistent with these prior estimates.^38–40^

### Minimum dispersion under scenarios without transmission by individuals with undetected infection

We first explored implications of the hypothesis that individuals who receive mpox diagnoses account for all transmission by estimating values of a dispersion parameter *k* that would correspond to this scenario. We measured the dispersion parameter as

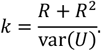

where *R* specified the reproduction number, or mean number of secondary infections (*U*) resulting from each index infection.^35–37^ Greater values of *k* correspond to lower degrees of dispersion in the number of infections caused by each index case; *k* approaches 0 as variance in *U* approaches ∞, and *k* approaches ∞ as variance in *U* approaches 0.

Maximum values of *k*, corresponding to the minimum degree of dispersion, arise under a scenario assuming uniform numbers of secondary infections caused by individuals with detected infections:

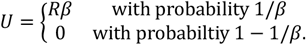

We sampled 10,000 distributions of *U* under each for *R* ∈ 0.5, 0.6, … 3.5, and repeated analyses under a scenario where only half of individuals at risk for transmitting infection received diagnoses (e.g., due to lack of clinical recognition of symptoms), where

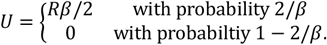

Across the range of *R* values considered, our analyses yielded maximum *k* estimates that remained consistently below the range of previously estimates,^38–40^ suggesting that the hypothesis of transmission only by diagnosed mpox cases implies an implausible degree of variation in individuals’ risk of transmitting MXPV.

### Contribution of undetected infections to transmission with realistic dispersion

We next sought to estimate the proportion of transmission that could be attributed to individuals with undetected infections under scenarios with *k*=0.3, consistent with published estimates.^38–40^ Defining *U*~Negative Binomial(*R* ∈ 0.7, 0.8, …, 3.5, *k* = 0.3), we drew values *U* and considered differing schemes for linking secondary infections to diagnosed or undiagnosed index cases.

Our first approach maximized the number of secondary infections attributed to diagnosed index cases. Defining *D* = *N*/*β* as the number of diagnosed cases among *N* infections, for a sorted vector *U*^′^ (with 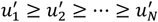), the proportion of infections attributable to diagnosed cases was 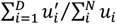, and the proportion attributable to undetected infections was 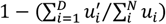.

To relax this deterministic sorting framework, we next considered scenarios where individuals’ likelihood of transmission was associated with being a diagnosed case. Defining *Y* as a vector indicating receipt of a diagnosis or no receipt of a diagnosis for a simulated population of cases (with probability 1/*β* of receiving a diagnosis), we defined 𝔼[*U*] = exp(*δ*_0_ + *δ*_1_*Y*) for *δ*_1_ values corresponding to 2-, 4-, 10-, and 20-fold greater numbers of secondary infections attributable to individuals diagnosed with mpox compared to undetected MPXV infections. We defined *δ*_0_ = ln[*R*/exp(*δ*_1_/*β*)] and solved via gradient descent for values of *θ* satisfying the condition

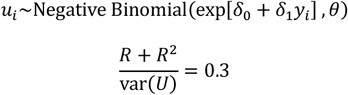

for sampled *Y* vectors of length 10,000.

Drawing from the resulting joint distributions of *Y* and *U*, the proportion of transmission attributable to individuals with diagnosed mpox was∑_*i*_ *y*_*i*_*u*_*i*_/∑_*i*_ *u*_*i*_, while the proportion attributable to individuals with undetected infections was ∑_*i*_(1 − *y*_*i*_)*u*_*i*_/∑_*i*_ *u*_*i*_.

### Elimination threshold assessment

Current WHO guidance defines mpox elimination in settings with surveillance capacity for confirmation of suspect cases, without zoonotic reservoirs for MPXV, and where outbreaks are concentrated in sexual networks, as ≥3 months without reported mpox cases.^7^ However, from a binomial distribution parameterized using our estimated reporting multiplier (1 in 33 [16-68]), the probability of observing *x*=0 diagnosed cases would be ≥50% with *n* equal to as many as 23 (11-47) true infections, and ≥5% with *n* as great as 98 (48-202) true infections. We used the modeling framework described above to estimate the probability that an outbreak was truly controlled under a scenario where ≥3 months had passed without additional notified cases.

We simulated transmission trees using a negative-binomial offspring distribution for values of *R* between 0.7-3.5 and *k*=0.3, using prior meta-analytic estimates of the serial interval distribution for clade IIb mpox in outbreaks driven by sexual transmission to draw inter-event times.^65^ We generated 100,000 simulated transmission chains for each value of *R*, and sampled cases receiving diagnoses among all simulated infections using our primary reporting multiplier estimate, halting simulations after either 2 years or the occurrence of >20,000 infections. We tabulated instances where 90 days passed after a notified case, with no other case notifications over this interval, and evaluated whether transmission chains had in fact concluded by assessing whether transmission chains included additional infections >90 months after the date of the last notified case. We stratified probabilities that transmission had concluded according to *R* and the total number of notified cases before onset of the monitoring period.

### Validation of under-reporting via meta-analysis of data from other settings

We conducted a systematic literature review of prior studies employing prospective testing for MPXV infection in anorectal specimens or anti-MPXV antibodies via blood specimens. We compared observed prevalence within the study populations to expected prevalence in the corresponding geographic areas based on local mpox case notification data from each setting, using a framework resembling our primary analyses within the KPSC study cohort. Analyses included corrections for potential differences in MPXV infection risk among individuals recruited in testing studies versus the general MSM population.

### Literature search

We searched PubMed for the terms (mpox OR monkeypox) AND (prevalen* OR inciden* OR surveill*) in full-length research articles published in English between 1 April, 2022 and 31 May, 2025, yielding 580 articles. We reviewed articles to identify those presenting results of studies that prospectively or retrospectively evaluated (*a*) prevalence of MPXV detection via anorectal samples among MSM who were not clinically suspected to be experiencing mpox disease, or (*b*) prevalence of anti-MPXV IgG detection among MSM who were not clinically suspected to be experiencing mpox disease, and who were not recruited based on clinical knowledge or suspicion of prior MPXV infection. We limited studies to those undertaken in regions where mpox was not understood to be endemic prior to 2022. Studies that did not exclude non-MSM from participation, or that did not present data stratified by MSM status, were eligible for inclusion if ≥70% of the sample were cisgender MSM or reported transgender/non-binary gender expression. Due to the rare nature of the outcome, we excluded studies that enrolled <100 individuals. To supplement our search, we conducted forward and backward citation tracking in Google Scholar from articles that met inclusion criteria to identify additional articles for screening.

To ensure comparability with our findings, we included results from anorectal specimens only when studies presented results from both anorectal and oropharyngeal specimen testing. We excluded studies that presented results from other specimen types (e.g., urine or saliva) without disaggregation of positive and negative results by specimen source. For serological studies that included vaccinated persons, we restricted eligibility to those using assays distinguishing naturally-acquired from MVA-induced antibody responses.

### Comparison of observed-to-expected prevalence in other settings

Within each setting where infection prevalence or seroprevalence studies were undertaken, we obtained publicly-reported mpox case counts at the finest-available temporal and spatial resolution. We estimated expected prevalence of infection, based on observed cases alone, by sampling dates of exposure (*τ*_*E*_), onset of MPXV shedding (*τ*_*S*_), and cessation of shedding (*τ*_*C*_), as described above for analyses within the KSPC study cohort; for settings where cases were aggregated by dates of reporting rather than dates of testing, we assumed a 1-day lag from the date a diagnostic test was performed to the reporting date. Where location-specific data were available on the proportion of mpox cases occurring among MSM, we multiplied daily case counts by this proportion to account for cases among MSM only; where such data were not available, we assumed MSM accounted for 95% of cases.^1^ For settings aggregating case notifications by week, we modeled daily diagnoses over the corresponding weeks as 1/7^th^ of the weekly total.

We estimated expected daily infection prevalence, accounting for reported cases alone, by dividing the total number of individuals expected to shed MPXV each day by estimates of the corresponding MSM population size. We obtained MSM population denominators for each geographic area via a literature search, using census data from each region to project changes in population size from historical estimates (**Table S8**). For serological studies, we assumed onset of detectable IgG at time of shedding cessation, based on previous evidence of times to seroresponse after natural infection^66,67^ and vaccination.^68^ We propagated uncertainty across 100,000 sampled time series of the daily prevalence of infection and seropositivity, and defined expected period-wide prevalence or seroprevalence as the mean of daily values over the enrollment period of each study, as described above for analyses comparing observed to expected prevalence within the KPSC study cohort.

### Risk normalization for reporting multipliers in other settings

Consistent with the rationale for inverse propensity weighting in the KPSC study, we expected that comparisons of MPXV infection prevalence or seroprevalence within study populations to mpox incidence among MSM in the surrounding geographic area could be biased by differences in risk among MSM enrolled in clinic-based samples and general-population samples. We used a two-stage procedure to correct for such bias. Because prior studies have provided estimates of the association of mpox risk with MSM’s number of anal sex partnerships over the preceding 21 days,^2,69^ and because total numbers of anal sex partnerships over specified intervals are a frequently-collected and frequently-reported characteristic of participants in sexual health research, we sought to reconstruct distributions of (*a*) anal sex partnerships per 3-week interval among MSM in clinic-based and general-population samples, and (*b*) relative risks of mpox, based on these sampled anal sex partnership counts.

We identified four studies reporting on anal sex partnership counts among MSM recruited in sexual health facilities;^70–72^ we constrained our search for studies to those undertaken within the US or Western Europe, as all included studies of MPXV prevalence were undertaken in these settings. We considered the sample enrolled in the ARTnet study as a reference population^73,74^ between July 2017 and January 2019. The sample for this study was recruited online among MSM directly after participation in the American Men’s Internet Study,^75^ an annual online behavioral survey recruiting MSM via email blasts and banner advertisements on websites or mobile phone applications targeting differing audiences (gay social networking, gay general interest, general social networking, and geospatial social networking). While the lack of a reference frame precludes comparison of the characteristics of the surveyed population to all MSM, consistency of STIs incidence within this study sample and US surveillance data suggest risk characteristics of the population are representative of those of MSM in general. Specifically, HIV prevalence within the study sample was 10.8%, comparable to the estimate of 11.1% prevalence of diagnosed HIV infection among all US MSM.^76^ Gonorrhea incidence within the sample was 5.07 cases/100 person-years, comparable to the estimate of 5.17 cases/100 person-years among MSM.^77^ Additionally, prior comparative studies have reported that sexual risk behaviors are similar in MSM samples recruited through such online surveys and those recruited via physical venue-based sampling.^78^

To account for the right-tailed shape of the distribution of individuals’ reported anal sex partnerships, we considered this count to follow a negative binomial distribution for the recall interval (*τ* days) used in each study, *Z*_*τ*_~Negative Binomial(*p, r*). We parameterized negative binomial distributions using either reported means and variances of distributions or via least-squares fitting to reported distribution quantiles. We sampled from the fitted distributions for pseudo populations of 1 million individuals and projected total partnership counts for individuals over a 3-week interval by multiplying the resulting samples by the factor 21/*τ*. To accommodate non-integer valued results, we drew counts probabilistically based on the remainder, with 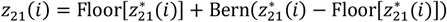. We used the association of anal sex partnership counts over a 21-day period from a test-negative design case-control study of mpox as the primary basis for assigning risk based on partnership counts.^2^ Compared to individuals who reported 0-1 anal sex partners in the 21 days preceding symptoms, adjusted odds ratios associated with reporting 2-3 partners and ≥4 partners were 2.2 (1.0-4.8) and 3.8 (1.7-8.8). We used these estimates to define and sample from probability distributions for individuals’ relative risk of mpox *π*(*i*) given their sampled partnership history, *f*(*π*(*i*)|*z*_21_(*i*)), considering the adjusted odds ratio and relative risk to be equivalent under the test-negative design framework.^79^ We repeated this analysis using alternative, model-based estimates of the relative risk of mpox given partnership history,^69^ which yielded similar risk multipliers for adjustment.

### Pooling of reporting reporting multipliers

We pooled estimates of the (log-transformed) reporting multiplier across all validation samples (molecular surveillance studies; serosurveillance studies) using inverse variance-weighted random effects models,^80^ for both unadjusted and risk-adjusted analyses (**Table S9**).

### Validation of under-reporting via phylogenetic analyses: overview

Last, we sought to estimate reporting multipliers within Los Angeles County by comparing the number of diagnosed cases to an estimate of the total MPXV viral population size generated by phylogenetic analyses. As detailed below, our workflow comprised generating a maximum-likelihood phylogeny of MPXV, generating estimates of the sampling proportion (*ρ*) accounting for both local transmission and repeated introductions of MPXV into Los Angeles County, and deriving the reporting multiplier *β* from our estimates of *p*. Here, *p* is defined as the ratio of the number of sequenced mpox cases to the effective population size (*N*_sequenced_/*N*_eff_) of MPXV in Los Angeles County (*38, 74, 75*). We describe these steps below together with a simulation study validating the framework for estimating *ρ*.

### Phylogenetic inference

We used all available MPXV clade IIb genomes found on Genbank on January 20, 2025 and identified local transmission clusters in Los Angeles County under a framework outlined in a previous study of MXPV phylodynamics.^39^ We created a temporally-resolved phylogeny using a modified version of the Nextstrain^83^ mpox workflow (https://github.com/nextstrain/mpox), which aligns sequences against the MK783032 (collection date: Nov. 2017) reference using nextalign.^84^ Briefly, this workflow included inferring a maximum-likelihood phylogeny using IQ-TREE^85^ with a general time-reversible nucleotide substitution model, estimating molecular clock branch lengths via TreeTime.^86^ The resulting phylogeny is available from https://nextstrain.org/groups/blab/mpox/allcladeIIseqs.

Our analyses included 497 de-duplicated MPXV sequences for Los Angeles County cases. Of this total, 271 were sampled between May 21, 2022 (the date of the first notified mpox case in California associated with the ongoing clade IIb outbreak) and October 14, 2022, and 226 were sampled between October 15, 2022 and December 31, 2024. The Los Angeles County Department of Public Health (LACDPH) sequences mpox cases diagnosed by healthcare providers in Los Angeles County, we assume that any genome sequenced by LACDPH was sampled locally. As the California Department of Health (CDPH) supports sequencing efforts only for local public health departments without internal sequencing capacity, we assumed additional MPXV sequences (*N*=222) sequenced by CDPH were sampled in locations within California outside of Los Angeles County. We clustered all Los Angeles County sequences based on inferred internal node location using a parsimony-based approach to reconstruct the locations of internal nodes.^39,51^

We inferred the time-varying effective reproduction number (*R*_*t*_), *ρ*, and phylogenies from all local clusters jointly using a birth-death skyline model,^48^ assuming that local outbreak clusters represented independent stochastic realizations of a transmission process with the same parameters.^87^ We assumed *R*_*t*_ to be piecewise-constant in intervals of 7 days, with

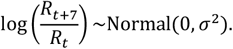

We estimated *σ* via Markov chain Monte Carlo sampling (MCMC) using BEAST2.^49^ We allowed an 8-day mean infectious period (*T*_g_), reflecting our assumption that detectable shedding of MPXV genetic material outlasts individuals’ period at risk of onward transmission (as described previously^22^ and accounted for in our estimates of expected infection prevalence in anorectal specimens (**Figure S2**)). We employed a strict molecular clock with a fixed value of 0.00006,^38^ and a HKY+Γ nucleotide substitution model with a fixed *κ* = 18.5.^88^ Of note, the birth-death skyline model conditions on survival,^48^ meaning that it computes the probability of observing a phylogenetic tree conditional on observing at least one lineage, and assumes the host population to be unstructured. Code for implementing this adapted version is available from https://github.com/nicfel/bdsky.

### Validation of estimation framework

Birth-death skyline models assume unstructured populations, and hidden population structure may bias inference results. Further, they do not model skewed offspring distributions as is the case for mpox transmission. To test the validity of our approach to estimating *ρ* under realistic mpox transmission dynamics, we simulated phylogenetic trees under using a stochastic, individual-based Susceptible-Exposed-Infectious-Recovered (SEIR) model with superspreading.^89^ We modeled importation assuming a constant force of introduction per unit time, and (as described above) assumed an offspring distribution with *U*~Negative Binomial(*R, k*). Primary analyses defined *k* = 0.3, consistent with previous estimates for MPXV;^38–40^ we additionally explored performance of the method with *k* = 0.1 (represent extreme dispersion) and *k* = 1 (representing low dispersion). To approximate real-life sampling dynamics, we parameterized time to sampling via the estimated time to healthcare presentation among UK mpox patients in 2022.^90^ We also tested performance under lower sampling schemes reducing the sampling by 50% and 90%, relative to observed levels. We recorded structured phylogenetic trees from model output and simulated corresponding genetic sequences using Seq-Gen,^91^ assuming an HKY substitution model, a genome size of 197,000bps, and a clock rate of 0.00006. We then ran simulated outputs through our multi-tree birth-death modeling pipeline and compared the estimated sampling proportion to input parameter values (**Figure S6**).

### Relating the sampling proportion to the number of infections

We used a previous derivation^50^ of the relationship between *N*_eff_ and the total number of infections, *N*_tot_, to generate reporting fractions based on our estimates of *ρ* from each analysis period (May 21 through October 14, 2022 and October 15, 2022 through December 31, 2024). Defining a coalescence rate *λ* = 1/(*N*_eff_*T*_g_) and an offspring distribution *U*~Negative Binomial(*R, k*),

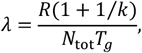

such that *N*_tot_ = *R*(1 + 1/*k*)*N*_eff_. With *ρ* = *N*_sequenced_/*N*_eff_, we obtain numerical estimates of *N*_tot_ during each analysis period as

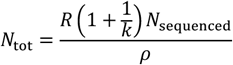

and obtain conditional values of *β*_*R*_ by dividing total diagnosed cases by *N*_tot_. We present estimates of *β*_*R*_ across a range of *R* estimates. Additionally, we obtain period-specific pooled estimates for *β* weighting these conditional values by Pr(*R_t_* = *R*) for mean *R_t_* values over each analysis period, i.e.

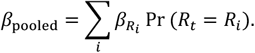

## Data Availability

Individuals wishing to access study data must enter into a data access agreement with Kaiser Permanente Southern California.

## Acknowledgments

The authors are indebted to the staff of the Molecular Microbiology Department at the Southern California Permanente Medical Group, Regional Reference Laboratory, Chino Hills for their support and contributions to this study.

## Funding

This work was funded by the Center for Forecasting and Outbreak Analytics of the US Centers for Disease Control & Prevention via the Insight Net cooperative agreement (CDC-RFA-FT-23-0069 to SYT and JAL). Its contents are solely the responsibility of the authors and do not necessarily represent the official views of the Centers for Disease Control and Prevention.

## Author contributions

Conceptualization: JAL, MIP, NFM, SYT; Methodology: JAL, MIP, GSD, UP, NFM; Investigation: JAL, MIP, MY, GSD, VH, JS, UP, NTP, LCG, NFM, SYT; Visualization: JAL, MIP; Funding acquisition: JAL, IRB, SYT; Project administration: JAL, MEP, IACR, SYT; Supervision: JAL, NFM, SYT; Writing—original draft: JAL, MIP, NFM; Writing—review & editing: JAL, MIP, MY, GSD, VH, JS, UP, NTP, LCG, MEP, IACR, IRB, NFM, SYT.

## Competing interests

The authors report no conflicts of interest relevant to this work.

## Data and materials availability

Analysis code is available via the corresponding author’s GitHub (joelewnard/mpox); individuals wishing to access to primary electronic health records data from Kaiser Permanente Southern California must enter into a signed agreement with the institution.

## Notes

### Competing Interest Statement

The authors have declared no competing interest.

### Author Declarations

The Kaiser Permanente Southern California Institutional Review Board approved this study with a waiver of informed consent for analysis of remnant specimens and patient electronic health records.

### Summary of Updates

-PCR testing controls and cycle threshold values -Analysis of elimination thresholds

