## Supporting information for "Extensive cryptic circulation sustains mpox among men who have sex with men"

**Contents of this supplement**

| Item | Title | Page |
|---|---|---|
| Table S1 | Characteristics of the study cohort and individuals tested. | 2 |
| Table S2 | Characteristics of individuals testing positive for MPXV in testing study and those receiving mpox diagnoses. | 4 |
| Table S3 | Assay results for positive specimens. | 5 |
| Table S4 | Under-reporting estimates in sensitivity analyses limiting eligible specimens for analyses. | 6 |
| Table S5 | Under-reporting estimates in sensitivity analyses with alternative natural history parameterization. | 7 |
| Table S6 | Estimated effectiveness of JYNNEOS vaccination. | 8 |
| Table S7 | Studies of MPXV infection prevalence or seroprevalence. | 9 |
| Table S8 | Analytic basis for estimation of MSM population sizes for meta-analysis of surveillance studies. | 10 |
| Table S9 | Pooled estimates of under-reporting multipliers across other studies. | 11 |
| Table S10 | Symptoms among individuals testing positive for MPXV in molecular surveillance studies. | 12 |
| Table S11 | Real-time PCR assay sequences. | 13 |
| Figure S1 | Characteristics of the KPSC study cohort. | 14 |
| Figure S2 | Modeled natural history of MPXV shedding in relation to date of reporting. | 15 |
| Figure S3 | Estimated reporting multipliers according to alternative test specificity values. | 16 |
| Figure S4 | Expected and observed MPXV infection prevalence or seroprevalence in other settings. | 17 |
| Figure S5 | Risk adjustment for comparison of individuals recruited in STI clinic settings to the general MSM population. | 18 |
| Figure S6 | Sensitivity of birth-death skyline model to sampling and transmission heterogeneity. | 19 |
| Figure S7 | Under-reporting estimates applying alternative estimates of dispersion. | 20 |
| Text S1 | Supplemental references | 21 |

**Table S1: Characteristics of the study cohort and individuals tested.**

| Characteristic | | Frequency, *n* (%) | | | Reweighted frequency[1] | | |
|---|---|---|---|---|---|---|---|
| | | Full cohort | Individuals tested | Individuals not tested | Full cohort | Individuals tested | Individuals not tested |
| | | *N*=7,930 | *N*=1,054 | *N*=6,876 | | | |
| Age group | | | | | | | |
| | 16-25 | 996 (12.6) | 70 (6.6) | 926 (13.5) | 12.5% | 11.9% | 12.6% |
| | 26-35 | 3,310 (41.7) | 460 (43.6) | 2,850 (41.4) | 41.9% | 43.0% | 41.7% |
| | 36-45 | 2,714 (34.2) | 407 (38.6) | 2,307 (33.6) | 33.9% | 31.9% | 34.2% |
| | ≥46 | 910 (11.5) | 117 (11.1) | 793 (11.5) | 11.7% | 13.2% | 11.5% |
| Race/ethnicity | | | | | | | |
| | White, non-Hispanic | 2,244 (28.3) | 319 (30.3) | 1,925 (28.0) | 28.1% | 27.1% | 28.3% |
| | Black, non-Hispanic | 738 (9.3) | 92 (8.7) | 646 (9.4) | 9.4% | 10.1% | 9.3% |
| | Asian/Pacific Islander, non-Hispanic | 797 (10.1) | 109 (10.3) | 688 (10.0) | 10.4% | 12.5% | 10.1% |
| | Hispanic, any race | 3,340 (42.1) | 417 (39.6) | 2,923 (42.5) | 42.0% | 41.1% | 42.1% |
| | Other/unknown | 811 (10.2) | 117 (11.1) | 694 (10.1) | 10.1% | 9.2% | 10.2% |
| Health insurance source | | | | | | | |
| | Commercial plan | 5,677 (71.6) | 760 (72.1) | 4,917 (71.5) | 71.4% | 70.3% | 71.6% |
| | Pre-paid/self-payment plan | 1,071 (13.5) | 162 (15.4) | 909 (13.2) | 13.4% | 12.5% | 13.5% |
| | Medicaid plan | 854 (10.8) | 85 (8.1) | 769 (11.2) | 11.1% | 12.9% | 10.8% |
| | Other/unknown | 328 (4.1) | 47 (4.5) | 281 (4.1) | 4.2% | 4.4% | 4.1% |
| Neighborhood deprivation index[2] | | | | | | | |
| | Quartile 1 (least deprived) | 2,525 (31.8) | 373 (35.4) | 2,152 (31.3) | 31.9% | 32.4% | 31.9% |
| | Quartile 2 | 1,817 (22.9) | 227 (21.5) | 1,590 (23.1) | 23.1% | 24.3% | 22.9% |
| | Quartile 3 | 1,647 (20.8) | 184 (17.5) | 1,463 (21.3) | 20.9% | 21.4% | 20.8% |
| | Quartile 4 (most deprived) | 1,934 (24.4) | 270 (25.6) | 1,664 (24.2) | 24.0% | 21.8% | 24.3% |
| | Unknown | 7 (0.1) | 0 (0.0) | 7 (0.1) | 0.1% | 0.0% | 0.1% |
| Prior-year healthcare utilization | | | | | | | |
| | <5 outpatient visits | 2,086 (26.3) | 174 (16.5) | 1,912 (27.8) | 26.4% | 27.3% | 26.3% |
| | 5-10 outpatient visits | 3,603 (45.4) | 571 (54.2) | 3,032 (44.1) | 45.3% | 44.1% | 45.4% |
| | 11-20 outpatient visits | 1,691 (21.3) | 238 (22.6) | 1,453 (21.1) | 21.3% | 21.2% | 21.3% |
| | ≥21 outpatient visits | 550 (6.9) | 71 (6.7) | 479 (7.0) | 7.0% | 7.4% | 6.9% |
| | Any emergency department visit | 1,376 (17.4) | 171 (16.2) | 1,205 (17.5) | 17.2% | 16.4% | 17.4% |
| | Any inpatient admission | 235 (3.0) | 26 (2.5) | 209 (3.0) | 2.9% | 2.3% | 3.0% |
| Prior-year gonorrhea testing | | | | | | | |
| | 0-1 tests | 2,132 (26.9) | 97 (9.2) | 2,035 (26.9) | 27.0% | 27.9% | 26.9% |
| | 2-5 tests | 4,967 (62.6) | 767 (72.8) | 4200 (61.1) | 62.6% | 62.3% | 62.6% |
| | ≥6 tests | 831 (10.5) | 190 (18.0) | 641 (9.3) | 10.4% | 9.8% | 10.5% |
| Gender and sexual orientation | | | | | | | |
| | Known male partners | 5,077 (64.0) | 815 (77.3) | 4,262 (62.0) | 63.3% | 58.8% | 64.0% |
| | Transgender identity | 268 (3.4) | 6 (0.6) | 262 (3.8) | 3.7% | 5.6% | 3.4% |
| Sexual health measures | | | | | | | |
| | HIV pre-exposure prophylaxis | 4,422 (55.8) | 831 (78.8) | 3,591 (52.2) | 55.4% | 53.1% | 55.7% |
| | Doxycycline post-exposure prophylaxis | 447 (5.6) | 69 (6.5) | 378 (5.5) | 5.8% | 6.8% | 5.6% |
| | 1 JYNNEOS dose | 1,231 (15.5) | 172 (16.3) | 1,059 (15.4) | 15.6% | 16.2% | 15.5% |
| | ≥2 JYNNEOS doses | 3,462 (43.7) | 631 (59.9) | 2,831 (41.2) | 43.8% | 45.0% | 43.7% |
| History of sexually transmitted infections | | | | | | | |
| | HIV infection | 1,435 (18.1) | 165 (15.7) | 1,270 (18.5) | 18.6% | 21.4% | 18.1% |
| | Prior syphilis diagnosis | 1,073 (13.5) | 160 (15.2) | 913 (13.3) | 13.9% | 16.0% | 13.5% |
| | Prior-year gonorrhea: 1 diagnosis | 965 (12.2) | 202 (19.2) | 763 (11.1) | 12.5% | 14.1% | 12.2% |
| | Prior-year gonorrhea: ≥2 diagnoses | 227 (2.9) | 52 (4.9) | 175 (2.5) | 2.9% | 2.8% | 2.9% |
| | Prior-year chlamydia: 1 diagnosis | 841 (10.6) | 156 (14.8) | 685 (10.0) | 10.8% | 12.0% | 10.6% |
| | Prior-year chlamydia: ≥2 diagnoses | 123 (1.6) | 31 (2.9) | 92 (1.3) | 1.6% | 1.8% | 1.6% |
| | Other STI in prior year | 21 (0.3) | 1 (0.1) | 20 (0.3) | 0.2% | 0.1% | 0.3% |
| | Prior mpox diagnosis | 9 (0.1) | 2 (0.2) | 7 (0.1) | 0.1% | 0.1% | 0.1% |
| Other risk factors | | | | | | | |

| | | | | | | |
|---|---|---|---|---|---|---|
| Alcohol or drug abuse diagnosis | 821 (10.4) | 93 (8.8) | 728 (10.6) | 10.3% | 9.7% | 10.4% |

[1]We calculate stabilized weights via logistic regression with receipt of testing as the outcome variable to balance characteristics of individuals who did or did not receive testing.
[2]Neighborhood deprivation index is computed at the census-tract level according to a previously-described framework.[1]

**Table S2: Characteristics of individuals testing positive for MPXV in testing study and those receiving mpox diagnoses.**

| Characteristic | | Frequency, *n* (%) | | |
|---|---|---|---|---|
| | | MPXV negative (testing study) | MPXV positive (testing study) | Diagnosed mpox cases |
| | | *N*=1,048 | *N*=6 | *N*=15 |
| Age group | | | | |
| | 16-25 | 70 (6.7) | 0 (0.0) | 1 (6.7) |
| | 26-35 | 457 (43.6) | 3 (50.0) | 8 (53.3) |
| | 36-45 | 405 (38.6) | 2 (33.3) | 5 (33.3) |
| | ≥46 | 116 (11.1) | 1 (16.7) | 1 (6.7) |
| Race/ethnicity | | | | |
| | White, non-Hispanic | 316 (30.2) | 3 (50.0) | 2 (13.3) |
| | Black, non-Hispanic | 92 (8.8) | 0 (0.0) | 1 (6.7) |
| | Asian/Pacific Islander, non-Hispanic | 108 (10.3) | 1 (16.7) | 1 (6.7) |
| | Hispanic, any race | 415 (39.6) | 2 (33.3) | 10 (66.7) |
| | Other/unknown | 117 (11.2) | 0 (0.0) | 1 (6.7) |
| Health insurance source | | | | |
| | Commercial plan | 756 (72.1) | 4 (66.7) | 13 (86.7) |
| | Pre-paid plan | 162 (15.5) | 0 (0.0) | 1 (6.7) |
| | Medicaid plan | 85 (8.1) | 0 (0.0) | 1 (6.7) |
| | Other/unknown | 45 (4.3) | 2 (33.3) | 0 (0.0) |
| Neighborhood deprivation index[1] | | | | |
| | Quartile 1 (least deprived) | 373 (35.6) | 0 (0.0) | 3 (20.0) |
| | Quartile 2 | 225 (21.5) | 2 (33.3) | 5 (33.3) |
| | Quartile 3 | 183 (17.5) | 1 (16.7) | 1 (6.7) |
| | Quartile 4 (most deprived) | 267 (25.5) | 3 (50.0) | 6 (40.0) |
| Prior-year healthcare utilization | | | | |
| | <5 outpatient visits | 173 (16.5) | 1 (16.7) | 2 (13.3) |
| | 5-10 outpatient visits | 569 (54.3) | 2 (33.3) | 11 (73.3) |
| | 11-20 outpatient visits | 235 (22.4) | 3 (50.0) | 2 (13.3) |
| | ≥21 outpatient visits | 71 (6.8) | 0 (0.0) | 0 (0.0) |
| | Any emergency department visit | 169 (16.1) | 2 (33.3) | 4 (26.7) |
| | Any inpatient admission | 26 (2.5) | 0 (0.0) | 1 (6.7) |
| Prior-year gonorrhea testing | | | | |
| | 0-1 tests | 96 (9.2) | 1 (16.7) | 1 (6.7) |
| | 2-5 tests | 764 (72.9) | 3 (50.0) | 13 (86.7) |
| | ≥6 tests | 188 (17.9) | 2 (33.3) | 1 (6.7) |
| Gender and sexual orientation | | | | |
| | Known male partners | 809 (77.2) | 6 (100.0) | 11 (73.3) |
| | Transgender identity | 6 (0.6) | 0 (0.0) | 1 (6.7) |
| Sexual health measures | | | | |
| | HIV pre-exposure prophylaxis | 827 (78.9) | 4 (66.7) | 10 (66.7) |
| | Doxycycline post-exposure prophylaxis | 68 (6.5) | 1 (6.7) | 2 (13.3) |
| | 1 JYNNEOS dose | 170 (16.2) | 2 (33.3) | 3 (20.0) |
| | ≥2 JYNNEOS doses | 628 (59.9) | 3 (50.0) | 4 (26.7) |
| History of sexually transmitted infections | | | | |
| | HIV infection | 164 (15.6) | 1 (16.7) | 3 (20.0) |
| | Prior syphilis diagnosis | 158 (15.1) | 2 (33.3) | 4 (26.7) |
| | Prior-year gonorrhea: 1 diagnosis | 200 (19.1) | 2 (33.3) | 3 (20.0) |
| | Prior-year gonorrhea: ≥2 diagnoses | 52 (5.0) | 0 (0.0) | 1 (6.7) |
| | Prior-year chlamydia: 1 diagnosis | 156 (14.9) | 0 (0.0) | 5 (33.3) |
| | Prior-year chlamydia: ≥2 diagnoses | 31 (3.0) | 0 (0.0) | 0 (0.0) |
| | Other STI in prior year | 1 (0.1) | 0 (0.0) | 0 (0.0) |
| | Prior mpox diagnosis | 2 (0.2) | 0 (0.0) | 0 (0.0) |
| Other risk factors | | | | |
| | Alcohol or drug abuse diagnosis | 93 (8.9) | 0 (0.0) | 2 (13.3) |

[1]Neighborhood deprivation index is computed at the census-tract level according to a previously-described framework.[1]

### Table S3: Assay results for positive specimens.

| Individual | Cycle threshold value | |
|---|---|---|
| | Mpox | Non-variola *Orthopoxvirus* |
| A | 36.3 | 37.2 |
| B | 34.3 | 34.8 |
| C | 24.2 | 24.7 |
| D | 33.8 | 34.7 |
| E | 15.6 | 16.2 |
| F (first specimen) | 21.2 | 21.8 |
| F (second specimen) | 26.8 | 27.5 |

We present cycle threshold ($c_T$) values for quantitative polymerase chain reaction tests for mpox-specific and non-variola *Orthopoxvirus* probes among positive specimens; we provide forward and reverse primer and probe sequences in **Table S11**.

### Table S4: Under-reporting estimates in sensitivity analyses limiting eligible specimens for analyses.

| Estimate | Weighted estimates (95% confidence interval)[1] | | | | | |
|---|---|---|---|---|---|---|
| | MPXV infection prevalence (95% confidence interval), % | | MPXV infection incidence per 100 person-years at risk | | Under-reporting multiplier (95% confidence interval) | |
| | *Clinically undetected and diagnosed cases* | *Expected from diagnosed cases only*[2] | *Clinically undetected and diagnosed cases* | *Diagnosed cases only* | *Time-to-event analysis (primary)* | *Instantaneous prevalence comparison*[3] |
| All testing results included (*primary*) | 0.91 (0.40, 1.63) | 0.035% | 22.2 (8.4, 48.0) | 0.61 | 33.1 (16.2, 67.6) | 26.3 (11.6, 46.5) |
| Excluding repeat positive results | 0.86 (0.38, 1.57) | 0.035% | 21.1 (7.9, 46.3) | 0.61 | 31.4 (15.2, 65.2) | 25.1 (10.8, 45.4) |
| Excluding consecutive samples at <30-day intervals | 0.88 (0.39, 1.59) | 0.035% | 21.5 (8.0, 46.9) | 0.61 | 31.9 (15.4, 66.4) | 25.3 (10.9, 45.5) |

[1]We calculate stabilized weights via logistic regression with receipt of testing as the outcome variable to balance characteristics of individuals who did or did not receive testing (**Table S1**).

[2]We estimate expected prevalence, accounting for diagnosed cases alone, by projecting time series of the number of individuals shedding infection each day, based on sampled times of onset and cessation of shedding.

[3]We compare observed to expected prevalence throughout the study period according to the ratio $\beta = p/\bar{\pi}$.

**Table S5: Under-reporting estimates in sensitivity analyses with alternative natural history parameterization.**

| Estimate | Weighted estimates (95% confidence interval)[1] | |
|---|---|---|
| | MPXV infection incidence, clinically undetected and diagnosed cases, per 100 person-years at risk | Under-reporting multiplier, time-to-event framework (95% confidence interval) |
| Parameterizing anorectal shedding duration from cross-sectional viral load measurements[1] | 14.3 (5.5, 30.7) | 21.3 (10.5, 42.9) |
| Parameterizing anorectal shedding duration from baseline and follow-up anorectal specimen testing[2] | 34.7 (13.1, 75.2) | 51.8 (25.2, 105.9) |

[1]Estimates are based on a parameterization of time to loss of detectable anorectal shedding from a study reporting viral loads from anorectal specimens collected 1-20 days after symptoms onset.[2] We projected daily viral load distributions using a linear regression model fitted to the study data, and used these samples to define the cumulative distribution function for time to loss of detectable shedding.

[2]Estimates are based on a parameterization of time to loss of detectable anorectal shedding from a study[3] reporting results of MPXV detection in anorectal specimens collected at baseline and follow-up assessments, which occurred 1-8 and 21-32 days after symptoms onset, respectively.

**Table S6: Estimated effectiveness of JYNNEOS vaccination.**

| Measure | Framework | Description | Data for comparison groups | Estimate (95% confidence interval) |
|---|---|---|---|---|
| Vaccine effectiveness against diagnosed mpox, $\mathrm{VE}_D = (1 - \theta_S \theta_P) \times 100\%$ | Case-control design | Compares adjusted odds of prior vaccination among individuals with diagnosed mpox (cases) versus individuals with gonorrhea (controls) via conditional logistic regression. Matching strata included individuals' HIV infection status and (among HIV-negative individuals) receipt or non-receipt HIV PrEP; receipt of doxycycline pre-exposure prophylaxis; and history of any syphilis diagnosis. Models further controlled for individuals' age group, receipt of *N. gonorrhea*/*C. trachomatis* testing in the prior year, and commercial or non-commercial insurance source (expected to proxy socioeconomic status) via covariate adjustment. | <u>Controls (*gonorrhea diagnosed*)</u> | |
| | | | Any vaccination: 135 (31.6%) | – – |
| | | | 1 dose: 83 (19.4%) | – – |
| | | | 2 doses: 209 (48.9%) | – – |
| | | | <u>Cases (*mpox diagnosed*)</u> | |
| | | | Any vaccination: 8 (53.3%) | 71.5% (9.1%, 91.1%) |
| | | | 1 dose: 3 (20.0%) | 54.7% (–89.2%, 89.1%) |
| | | | 2 doses: 4 (26.7%) | 78.3% (16.8%, 94.3%) |
| | Negative control-corrected cohort design | Compares time to mpox diagnosis among JYNNEOS recipients to non-recipients, dividing the estimated adjusted hazard ratio by the adjusted estimated hazard ratio conveying the association of JYNNEOS vaccination with a negative-control outcome (gonorrhea), per Sanderson-Smith et al.[4] We estimated adjusted incidence rate ratios for each outcome via Poisson regression, controlling for HIV infection, receipt of HIV PrEP, receipt of doxycycline post-exposure prophylaxis, prior syphilis diagnosis, age group, prior-year gonorrhea testing, and commercial insurance source as covariates. | <u>VE against mpox (*negative control-uncorrected*)</u> | |
| | | | No vaccination: 0.8/100 PYAR | – – |
| | | | Any vaccination: 0.5/100 PYAR | 59.6% (–17.9%, 86.1%) |
| | | | 1 dose: 0.8/100 PYAR | 29.1% (–176.5%, 81.7%) |
| | | | 2 doses: 0.4/100 PYAR | 69.6% (–6.1%, 91.4%) |
| | | | <u>VE against gonorrhea (*negative control outcome*)</u> | |
| | | | No vaccination: 20.8/100 PYAR | – – |
| | | | Any vaccination: 31.7/100 PYAR | –19.3% (–42.0%, –0.5%) |
| | | | 1 dose: 30.1/100 PYAR | –16.9% (–47.3%, 7.2%) |
| | | | 2 doses: 32.0/100 PYAR | –20.8% (–45.0%, –0.6%) |
| | | | <u>VE against mpox (*negative control-corrected*)</u> | |
| | | | Any vaccination | 66.0% (–0.1%, 88.4%) |
| | | | 1 dose | 39.3% (–140.7%, 84.7%) |
| | | | 2 doses | 74.9% (10.8%, 92.9%) |
| Vaccine effectiveness against MPXV infection, $\mathrm{VE}_S = (1 - \theta_S) \times 100\%$ | Matched test-negative design | Compares adjusted odds of prior vaccination among individuals testing positive for MPXV infection in anorectal specimens (cases) versus individuals testing negative (controls). Matching strata included individuals' HIV infection status and (among HIV-negative individuals) receipt or non-receipt HIV PrEP; receipt of doxycycline pre-exposure prophylaxis; and history of any syphilis diagnosis. | <u>Controls (*testing negative*)</u> | |
| | | | No vaccination: 274 (23.2%) | – – |
| | | | Any vaccination: 909 (76.8%) | – – |
| | | | 1 dose: 184 (15.6%) | – – |
| | | | 2 doses: 725 (61.3%) | – – |
| | | | <u>Cases (*testing positive*)</u> | |
| | | | No vaccination: 1 (14.3%) | – – |
| | | | Any vaccination: 6 (85.7%) | 52.6% (–166.9%, 91.6%) |
| | | | 1 dose: 2 (28.6%) | 41.4% (–432.1%, 93.6%) |
| | | | 2 doses: 4 (57.1%) | 58.8% (–157.6%, 93.4%) |
| | Cohort design | Compares weighted incidence rates of MPXV infection among vaccinated individuals to unvaccinated individuals, corrected for estimated degree of under-reporting within each stratum. | No vaccination: 35.8/100 PYAR | – – |
| | | | Any vaccination: 11.7/100 PYAR | 50.7% (–91.8%, 94.5%) |
| Vaccine effectiveness against progression to diagnosed mpox, given MPXV infection, $\mathrm{VE}_P = (1 - \theta_P) \times 100\%$ | Model-based estimate | Obtained numerically, defining the risk (hazard, rate, …) ratio of diagnosed mpox associated with vaccination as the product of the risk (hazard, rate, …) ratio of infection, given vaccination, times the risk (hazard, rate, …) ratio of progression to disease, given infection, associated with vaccination. | – – | 41.0% (–244.6%, 89.9%) |

PYAR: Person-year at risk.

**Table S7: Studies of MPXV infection prevalence or seroprevalence.**

| Study type | Location | Citation[1] | Population sampled | Dates | Observed prevalence[2] | Expected prevalence (95% confidence interval) |
|---|---|---|---|---|---|---|
| Molecular testing | | | | | | |
| | Paris, France | Ferré et al., 2022[5] | MSM living with HIV or taking HIV pre-exposure prophylaxis and receiving routine screening for *C. trachomatis* and *N. gonorrhoeae*, with no suspicion of mpox illness | June 5 to July 11, 2022 | 6.5% (13/200) | 0.088% (0.036–0.163%) |
| | Antwerp, Belgium | De Baetselier et al., 2022[6] and van Dijck, et al.[7] | MSM living with HIV or taking HIV pre-exposure prophylaxis and receiving routine screening for *C. trachomatis* and *N. gonorrhoeae*, with no suspicion of mpox illness | | | |
| | | | | May 1-30, 2022 | 1.8% (4/224) | 0.005% (0.002–0.009%) |
| | | | | June 1-30, 2022 | 4.8% (7/146) | 0.017% (0.009–0.025%) |
| | | | | July 1 to 30 September, 2022 | 3.9% (7/181) | 0.012% (0.001–0.028%) |
| | Geneva and Zürich, Switzerland | Hampel et al., 2023[8] | MSM taking HIV pre-exposure prophylaxis and receiving routine screening for *C. trachomatis* and *N. gonorrhoeae*, with no suspicion of mpox illness | August 23 to October 4, 2022 | 1.0% (2/201) | 0.006% (0.001–0.015%) |
| | Washington, DC, United States | Ogale et al., 2023[9] | Individuals presenting for their first JYNNEOS vaccine dose, with no suspicion of mpox illness | August 11-31, 2022 | | 0.031% (0.017–0.050%) |
| | | | *With restriction to anorectal specimens* | | 1.2% (2/164) | |
| | | | *Without restriction to anorectal specimens* | | 1.8% (3/164) | |
| | Barcelona, Spain | Agustí et al., 2024[10] | MSM recruited at a community center with no suspicion of mpox illness | August 1 to October 31, 2022 | | 0.024% (0.001–0.091%) |
| | | | *With restriction to anorectal specimens* | | 1.8% (2/112) | |
| | | | *Without restriction to anorectal specimens* | | 6.2% (7/113) | |
| Serological testing | | | | | | |
| | San Francisco, United States | Minhaj et al., 2023[11] | MSM receiving services at sexual health clinics with no suspicion of mpox illness | June 28 to August 26, 2022 | 8.0% (18/225) | 1.02% (0.96–1.09%) |
| | New York, United States | Pathela et al., 2024[12] | MSM receiving routine syphilis and HIV testing, with no history of receipt of vaccina virus-based vaccines and with no suspicion of prior mpox illness or exposure | July 7 to September 1, 2022 | 7.8% (16/166) | 1.32% (1.09–1.63%) |
| | Rome, Italy | Matusali et al., 2023[13] | Men presenting for their first JYNNEOS vaccine dose, with no history of receipt of vaccinia virus-based vaccines and with no prior mpox diagnosis | August 8 to September 9, 2022 | 12.8% (18/141) | 0.19% (0.16–0.23%) |
| | Washington, DC, United States | Ogale et al., 2023[9] | Individuals presenting for their first JYNNEOS vaccine dose, with no suspicion of mpox illness (limited to individuals providing anorectal swab) | August 11-31, 2022 | 14.5% (47/324) | 1.17% (0.97–1.44%) |
| | Berlin, Germany | Marcus et al., 2024[14] | MSM recruited in private practices and community-based checkpoints specialized in care for HIV and sexually-transmitted infections | April 1 to June 30, 2023 | 16.4% (149/908) | 2.36% (1.95–2.90%) |

[1]We identified studies through a systematic review including a PubMed search augmented by forward- and back-tracking of citations.
[2]Analyses restricted results from molecular testing studies to those generated from anorectal specimens. We restricted results from serological testing studies to those enrolling unvaccinated or previously uninfected individuals (for studies during early phases of the outbreak in summer, 2022), or those using an antibody marker that distinguishes natural infection from vaccine response (for studies undertaken after vaccine implementation).

### Table S8: Analytic basis for estimation of MSM population sizes for meta-analysis of surveillance studies.

| Setting | Estimate (95% CI) | Description |
|---|---|---|
| Île-de-France (Paris region), France | 95,661 (58,240–133,087) | A previous study[15] estimated the population of MSM within Île-de-France as of 2014 (*N*=93,178). We scaled this estimate by the population size of Île-de-France in 2014 and extrapolated to the population as of 2022. |
| Belgium | 111,620 (92,413–130,811) | Two prior estimates[16,17] of the population of MSM within Belgium were available (*N*=106,336 in 2013 and *N*=144,753 in 2015). We scaled these estimates by the population sizes of Belgium as of 2013 and 2015 and extrapolated to the population as of 2022. We multiplied by the population size of Belgium as of 2022 and assumed a normal distribution in pooling the estimates. |
| Lazio, Italy | 128,865 (105,388–152,348) | Direct estimates of the population size of MSM in Lazio or Rome, Italy were not available. A previous study[18] estimated that MSM accounted for 2.7% of the adult male population in Verona, Italy as of 2014 (*N*=10,952); we scaled this estimate by the size of the population of men aged 16-64 years in Verona and multiplied by the size of the population of adult men aged 16-64 in the metropolitan city of Rome to obtain the urban MSM population. We added 1% of the population size of men aged 16-64 within the remaining administrative districts within Lazio to account for MSM outside the urban core, assuming a lower proportion in such regions would be MSM. We assumed the same ratio of the mean to standard deviation as reported in Verona and assumed a normal distribution. |
| Switzerland | 88,758 (70,986–106,535) | A previous study[19] estimated the population of MSM within Switzerland as of 2011 (*N*=80,000). We scaled this estimate by the population size of Switzerland as of 2011, and multiplied by the population size of Switzerland as of 2022. We assumed a normal distribution. |
| Berlin, Germany | 94,931 (76,312 - 113,496) | A previous study[20] estimated the population of MSM within Berlin as of 2008 (*N*=91,000). We scaled this estimate by the population size of Berlin in 2008 and extrapolated to the population as of 2024. Measures of uncertainty were not provided with the original report. We therefore defined standard deviation in the estimate as being equal to 10% of the mean and assumed a normal distribution. |
| Barcelona, Spain | 42,613 (34,221 - 50,980) | A precious study[21] estimated the population of MSM comprised 5.3% of Barcelona's male population in 2012. Assuming this proportion remains constant through 2024, we extrapolated to the male population as of 2024. Measures of uncertainty were not provided with the original report. We therefore defined standard deviation in the estimate as being equal to 10% of the mean and assumed a normal distribution. |
| Washington, DC, United States | 34,857 (28,840–42,895) | A previous study[22] estimated the population of MSM within Washington, DC as of 2016 (*N*=35,867). We scaled this estimate by the population size of Washington, DC in 2016 and extrapolated to the population as of 2022. Measures of uncertainty were not provided with the original report. We therefore defined standard deviation in the estimate as being equal to 10% of the mean and assumed a normal distribution. |
| San Francisco, California, United States | 64,665 (60,607–68,730) | A previous study[23] estimated the population size of MSM within San Francisco as of 2017 (*N*=69,974). We scaled this estimate by the population size of San Francisco in 2017 and extrapolated to the population as of 2022. We assumed a normal distribution for reported uncertainty bounds and for our projection. |
| New York, New York, United States | 218,375 (175,551–261,181) | A previous study[22] estimated the population size of MSM within New York City as of 2016 (*N*=224,020). We scaled this estimate by the population size of New York City in 2016 extrapolated to the population as of 2022. Measures of uncertainty were not provided with the original report. We therefore defined standard deviation in the estimate as being equal to 10% of the mean and assumed a normal distribution. |

### Table S9: Pooled estimates of under-reporting multipliers across other studies.

| Studies included | Estimate (95% confidence interval)[1] | |
|---|---|---|
| | Without risk adjustment | With risk adjustment |
| All molecular testing studies | 189 (90.7, 394) | 136 (65.1, 283) |
| All serological testing studies | 12.9 (5.59, 29.7) | 9.24 (4.01, 21.3) |
| All molecular and serological testing studies | 54.5 (20.7, 143) | 39.1 (14.9, 103) |

[1]Pooled estimates are generated via inverse variance-weighted random effects models including under-reporting multipliers from each selection of studies.

**Table S10: Symptoms among individuals testing positive for MPXV in molecular surveillance studies.**

| Reference | Setting and design | Outcomes[1] | |
|---|---|---|---|
| | | Individuals followed | Individuals with symptoms or mpox diagnosis |
| Agustí et al., *Nat Comm* 2024[10] | Testing via self-collected oropharyngeal and anorectal specimens with telephone follow-up 21 days after any positive test for symptom interview | 7 | 2 |
| de Batselier et al., *Nat Med* 2022;[6] van Dijck et al., *Lancet Microbe* 2023[7] | Testing via oropharyngeal and anorectal specimens with accompanying symptom interview and clinical assessment, with follow-up 21-37 days after testing | 18 | 5 |
| Hampel et al., *Lancet Microbe* 2023[8] | Testing via oropharyngeal and anorectal specimens within an existing longitudinal cohort study collecting symptoms data via web application | 2 | 1 |
| Ferré et al., *Ann Intern Med* 2022[5] | Testing via anorectal specimens with subsequent outreach to individuals testing positive to collect symptoms data | 13 | 2 |
| Ogale et al., *Clin Infect Dis* 2024[9] | Testing via anorectal and pharyngeal specimens within a cohort study involving 3 clinic visits over two months with symptoms interview at study visits. | 3 | 1 |
| Kaiser Permanente Southern California cohort (current study) | Testing of anorectal specimens collected for STI screening, with monitoring for mpox-related healthcare utilization among cohort members in integrated healthcare system | 6 | 0 |
| **Total** | | 49 | 11 |

[1]Data encompass all individuals reported to experience infection, as detected by either oropharyngeal or anorectal specimens (a broader population than those included in analyses as reported in **Table S7**). Studies differed in monitoring for symptoms based on active or passive surveillance modalities.

**Table S11: Real-time PCR assay sequences.**

| Assay | Primer/probe | Sequence |
|---|---|---|
| Non-variola *Orthopoxvirus* | | |
| | Forward | 5'-TCA ACT GAA AAG GCC ATC TAT mGA-3' |
| | Reverse | 5'-GAG TAT AGA GCA CTA TTT CTA AAT CCmC A-3' |
| | Probe | 5' - /5YakYel/CCA TGC AAT/ZEN ATA CGT ACA AGA TAG TAG CCA AC / 3IABkFQ/-3' |
| Mpox | | |
| | Forward | 5'-ACG TGT TAA ACA ATG GGT GAmU G-3' |
| | Reverse | 5'-AAC ATT TCC ATG AAT CGT AGT mCC-3' |
| | Probe | 5'-/56-FAM/TGA ATG AAT /ZEN/ GCG ATA CTG TAT GTG TGG G/3IABkFG/-3' |

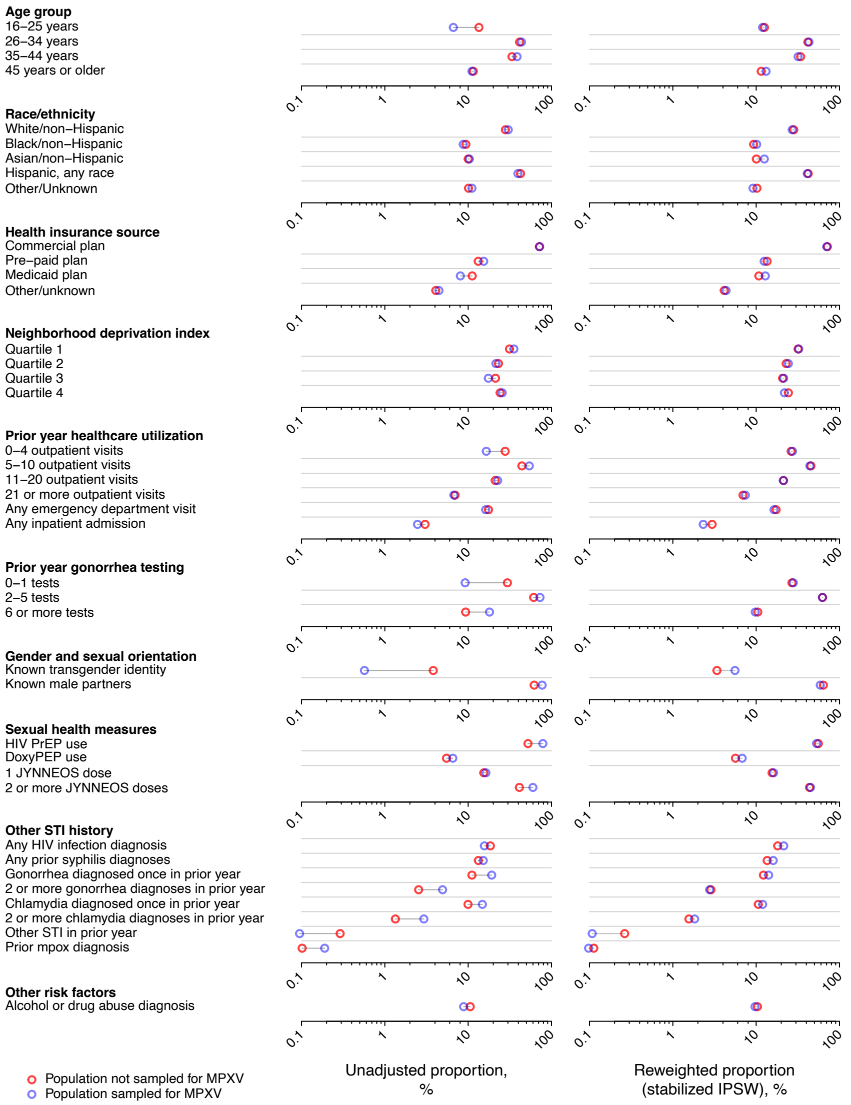

**Figure S1: Characteristics of the KPSC study cohort.** We present distributions of demographic and clinical characteristics within the (left) unweighted and (right) reweighted study cohort, with weights estimated by logistic regression to balance characteristics of individuals who received or did not receive MPXV testing by anorectal swabs. We present underlying data in **Table S1** and compare individuals testing positive or negative for MPXV, in the unweighted and weighted samples, in **Table S2**.

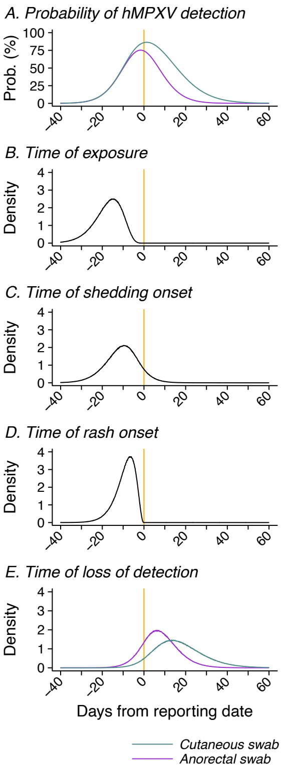

**Figure S2: Modeled natural history of MPXV shedding in relation to date of reporting.** We present (**A**) daily probabilities of MPXV detection in cutaneous and anorectal swabs, along with (**B**-**E**) fitted distributions for times-to-event for underlying epidemiologic parameters including times from exposure, onset of MPXV shedding, onset of clinical rash, and time to cessation of shedding detectable by PCR.

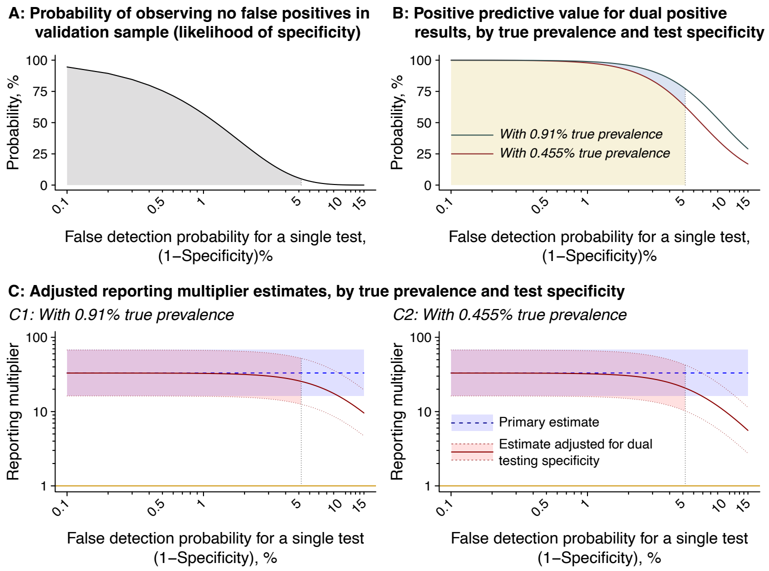

**Figure S3: Estimated reporting multipliers according to alternative test specificity values.** We present (**A**) the likelihood of the indicated single-test specificity values, given the observation of 0 false positives among 64 true negative specimens in an external assessment of the study assay.[24] We compute this value via a binomial distribution with $x$=0, $n$=64, and $p$ equal to the false detection probability for a single test (1–Specificity). The shaded area (for panel **A** and all subsequent panels) delineates the range of single-test false detection probabilities for which the probability of observing $x$=0 false positive results among $n$=64 specimens exceeds 95%. Next, we illustrate (**B**) estimates of positive predictive value for dual testing with true infection prevalence equal to 0.91% (as estimated in the study; blue) and 0.455% (half the estimated value; brown). Below, we illustrate (**C**) reporting multipliers for adjusted for potential false positives under the dual testing procedure. Red shaded areas delineate 95% confidence intervals around point estimates (center line; red). We illustrate primary estimates without specificity adjustment in blue (shaded area, 95% confidence interval; point estimate, dotted line). Panel A1 (left) illustrates reporting multipliers under an assumed true prevalence of 0.91%, and panel A2 (right) illustrates reporting multipliers under an assumed true prevalence of 0.455%.

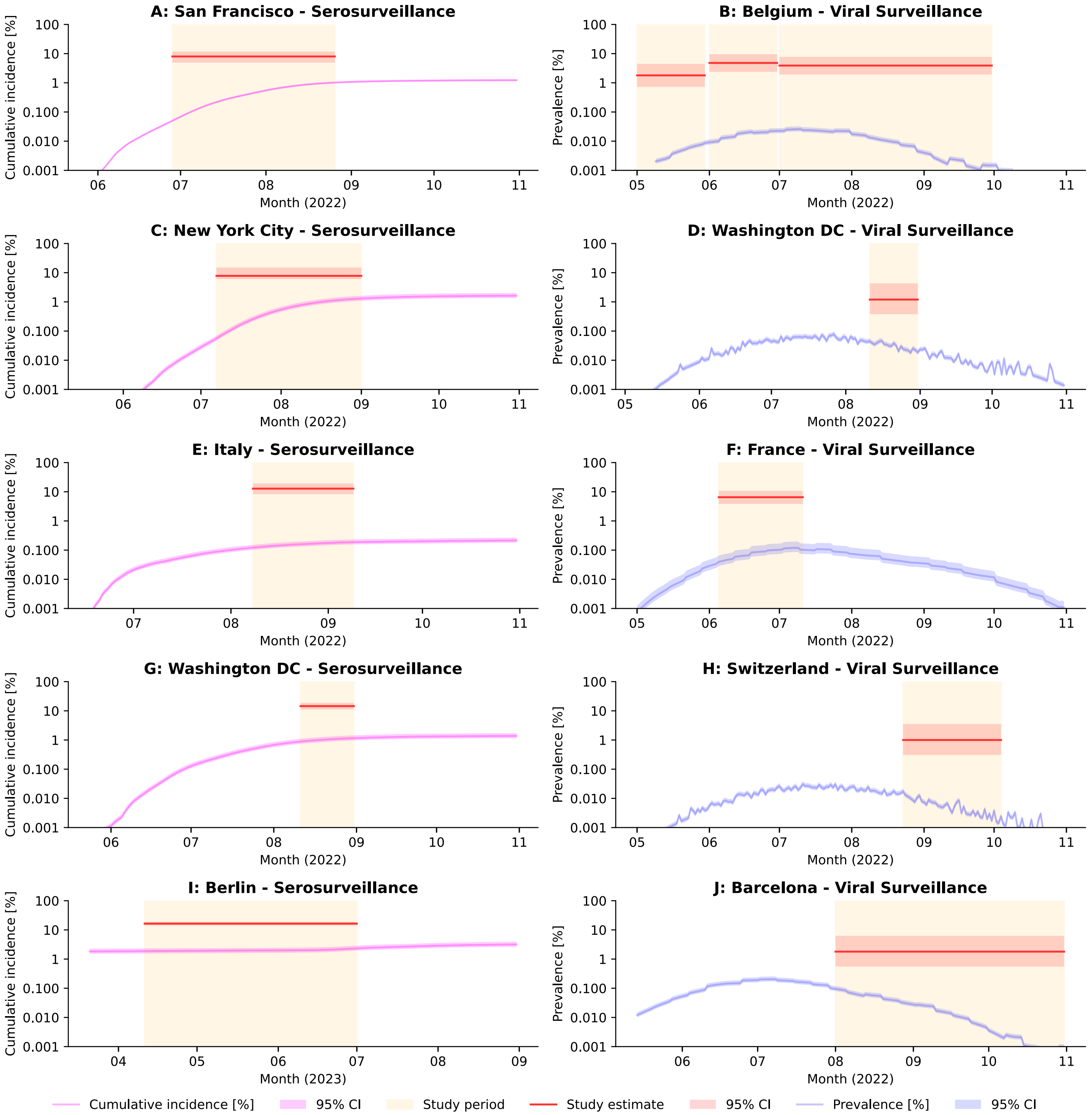

**Figure S4: Expected and observed MPXV infection prevalence or seroprevalence in other settings.** We illustrate estimated seroprevalence (left column; pink line, with shaded area delineating 95% confidence intervals) and infection prevalence (right column; blue line, with shaded area delineating 95% confidence interval) based on reported cases within each setting (**Table S6**; **Table S7**). Tan shaded areas delineate the study periods, with red lines denoting observations within each study (shaded areas delineate 95% confidence intervals around studies' prevalence or seroprevalence estimates).

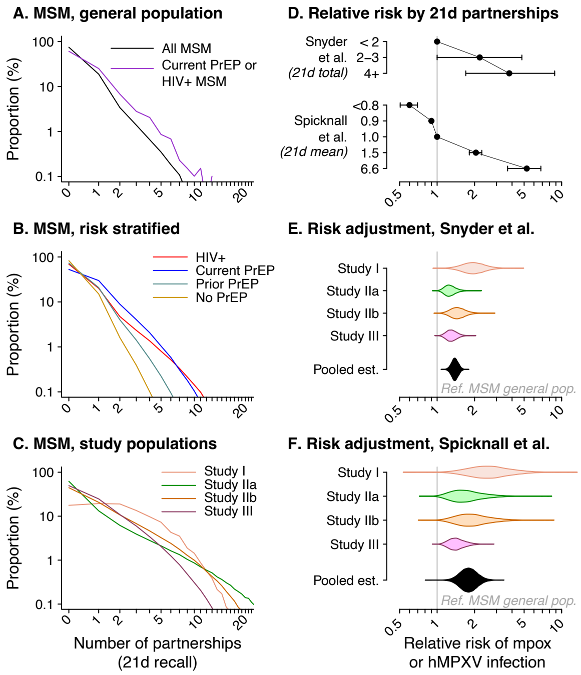

**Figure S5: Risk adjustment for comparison of individuals recruited in STI clinic settings to the general MSM population.** We present comparisons of partnership counts within a 21-day recall period among differing MSM strata within a general population sample (**A**, **B**), as reported in the ARTnet study,[25,26] along with the strongly right-tailed distribution of partnership counts within the same recall period among individuals recruited in sexual health facilities (**C**) based on three studies.[27–29] In the right-hand column, we illustrate reported associations of number of unprotected anal intercourse partnerships within a 21-day recall period with mpox risk based on a case-control study[30] and modeling study[31] (**D**). Below, we illustrate estimates of the relative risk of mpox among individuals recruited in sexual health facilities versus the general MSM population, for the individual studies presented in (**C**) and pooled across these studies, using relative risk parameters based on the case-control study (**E**) and the modeling study (**F**).

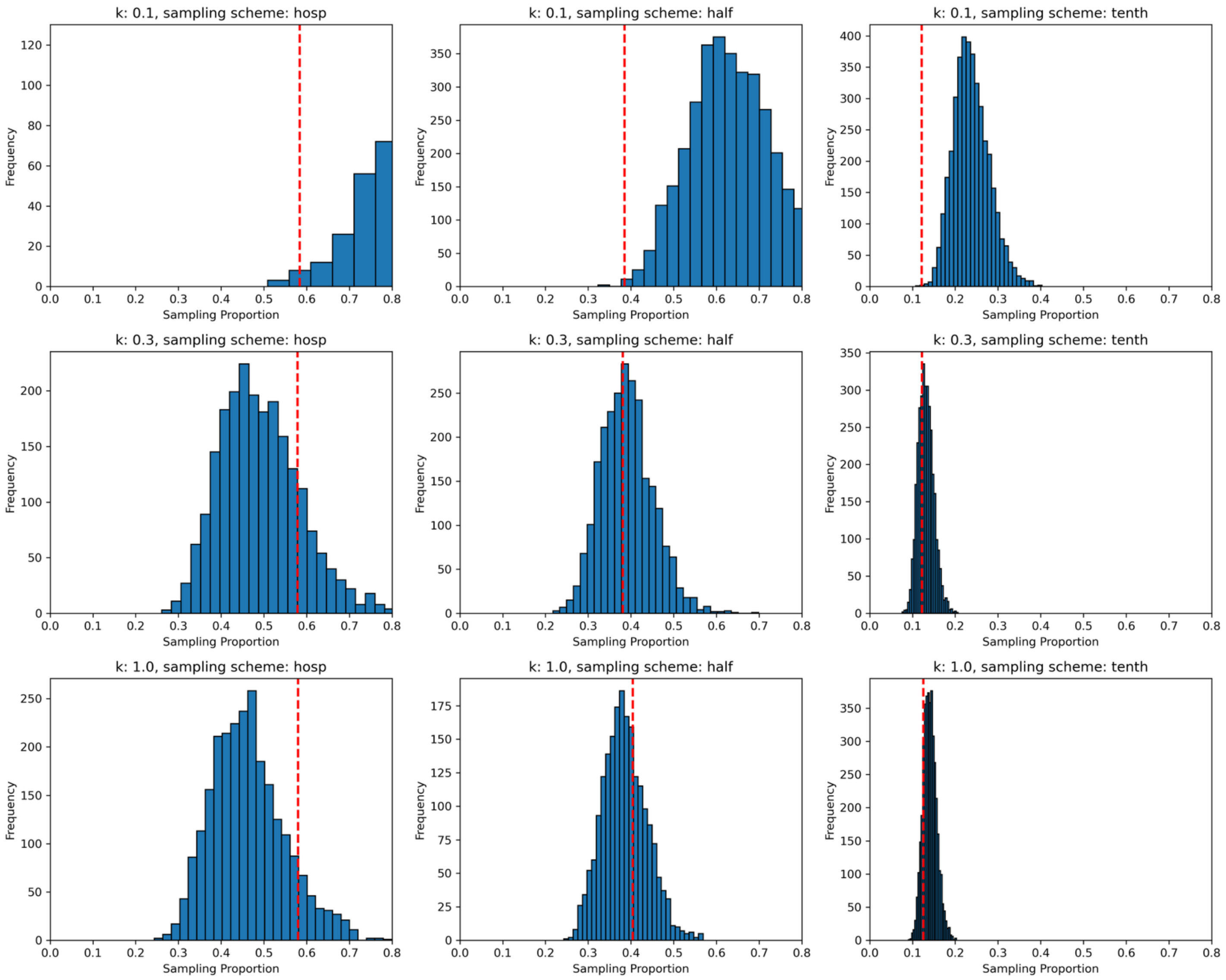

**Figure S6: Sensitivity of birth-death skyline model to sampling and transmission heterogeneity.** We illustrate the ability of our birth-death skyline model to accurately capture the true sampling proportions by simulating MPXV transmission dynamics with different values of the dispersion parameter (*k*=0.1, 0.3, 1.0) and sampling schemes. Under the sampling scheme "hosp", time to sampling is defined as the estimated time to present to healthcare in the UK in 2022;[32] the "half" and "tenth" sampling schemes reduce this rate by 50% and 90%, respectively. Histogram bars indicate the distribution of sampling proportion estimates from analyses of simulated data, while red dashed lines represent the true sampling proportion used to parameterize simulations. The center row of panels corresponds to $k = 0.3$, consistent with prior empirical findings,[33–35] and demonstrates the best alignment between true and estimated values of *p* under each sampling scheme.

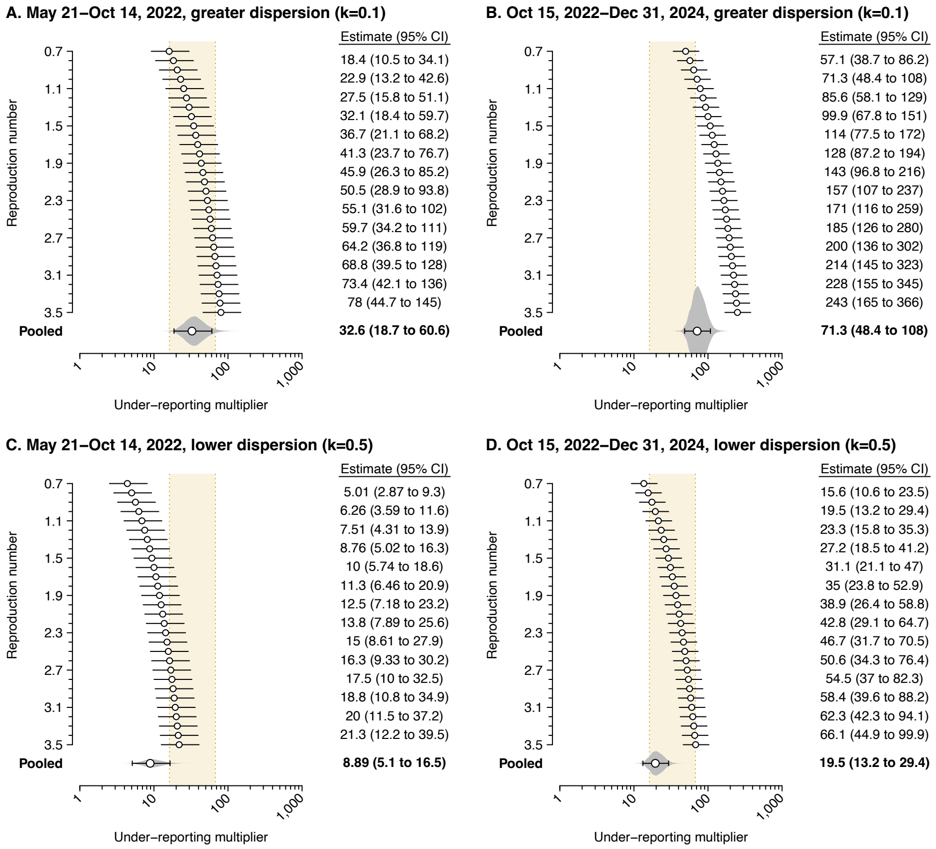

**Fig. S7: Under-reporting estimates applying alternative estimates of dispersion.** We illustrate under-reporting multipliers based on a comparison of diagnosed cases to the estimated population size of MPXV infections in Los Angeles County from phylogenetic analyses of sequenced cases, accounting for introductions from external geographies, for the periods from May 21 to October 14, 2022 and from October 15 to December 24, 2022. Within each panel, estimates are conditioned on the reproduction number, *R*. We illustrate the probability density of mean daily *R* values throughout the analysis period in purple. Pooled estimates presented in bold at the bottom of the figure are weighted according to the period-specific probability density of *R*. Top panels (**A**, **B**) present estimates under a scenario with extreme dispersion in the offspring distribution ($k = 0.1$), while bottom panels (**C**, **D**) consider a scenario with lower dispersion ($k = 0.5$).
